# Executable models of immune signaling pathways in HIV-associated atherosclerosis

**DOI:** 10.1101/2022.03.07.22271522

**Authors:** Mukta G. Palshikar, Rohith Palli, Alicia Tyrell, Sanjay Maggirwar, Giovanni Schifitto, Meera V. Singh, Juilee Thakar

## Abstract

Atherosclerosis (AS)-associated cardiovascular disease is an important cause of mortality in an aging population of people living with HIV (PLWH). This elevated risk has been attributed to viral infection, anti-retroviral therapy, chronic inflammation, and lifestyle factors. However, rates at which PLWH develop AS vary even after controlling for length of infection, treatment duration, and for lifestyle factors. To investigate the molecular signaling underlying this variation, we sequenced 9368 peripheral blood mononuclear cells (PBMCs) from eight PLWH, four of whom have atherosclerosis (AS+). Additionally, a publicly available dataset of PBMCs from persons before and after HIV infection was used to investigate the effect of acute HIV infection. To characterize dysregulation of pathways rather than just measuring enrichment, we developed the single-cell Boolean Omics Network Invariant Time Analysis (scBONITA) algorithm. scBONITA infers executable dynamic pathway models and performs perturbation analysis to identify high impact genes. These dynamic models are used for pathway analysis and to map sequenced cells to characteristic signaling states (attractor analysis). scBONITA revealed that lipid signaling regulates cell migration into the vascular endothelium in AS+ PLWH. Pathways implicated included AGE-RAGE and PI3K-AKT signaling in CD8+ T cells, and glucagon and cAMP signaling pathways in monocytes. Attractor analysis with scBONITA facilitated pathway-based characterization of cellular states in CD8+ T cells and monocytes. In this manner, we identify critical cell-type specific molecular mechanisms underlying HIV-associated atherosclerosis using a novel computational method.

## Introduction

Chronic human immunodeficiency virus (HIV) infection increases the risk of atherosclerosis (AS) associated cardiovascular disease (CVD), which is a leading cause of morbidity in persons living with HIV (PLWH) (1–4). PLWH have an increased prevalence of risk factors for AS (5–9), some of which are driven by the off-target effects of antiretroviral drugs (9, 10). However, even after controlling for these risk factors, the risk of CVD remains significantly higher in PLWH (4, 11, 12), suggesting a role of long-term HIV infection.

HIV infection causes metabolic changes leading to a pro-atherogenic inflammatory environment in the vasculature (13–16). HIV infection and long-term antiretroviral therapy mediate signaling dynamics, including inflammasome activation, cell migration and apoptosis, in PBMCs and the vasculature (17, 18). Biomarker studies highlight these atherogenic processes, especially in the context of activated monocyte/macrophages and T cells (14–16). CD8+ T cells contribute to atherogenesis by secretion of cytotoxic granules and formation of the necrotic core of atherosclerotic plaques (19). Monocyte/macrophages migrate into the intima and form apoptotic atherosclerotic plaques (20). CD4+ T cells and B cells have also been previously implicated in CVD (21–31). The interplay between signaling pathways, immune cell activation and inflammation in HIV infection requires further investigation. Single-cell sequencing (scRNA-seq) allows simultaneous investigation of these perturbations. To investigate the mechanistic link between HIV infection and atherosclerosis, we sequenced PBMCs from PLWH with and without atherosclerosis, matched for length of infection, cART, and other AS risk factors (Supplementary Note 1, Supplementary Figure 1).

Inference of mechanisms from high-dimensional scRNA-seq data is not trivial (32). Typically, scRNA-seq analysis uses clustering to define cell subpopulations, followed by differential expression (DE) and gene set overrepresentation analysis (ORA) to estimate pathway modulation. This approach discounts pathway topology and cannot connect molecular state to cellular state. ORA ignores synergistic interactions among genes by treating genes as independent and equal, and fails to estimate the significance of pathways (33). Our previously published algorithm overcomes above caveats by using discrete-state network modeling to perform pathway analysis using bulk transcriptomic data (34). This algorithm has been rigorously tested and compared to other pathway analysis methods (35). This algorithm employs discrete-state network modeling, which uses Boolean rules to explicitly define signal integration. These rules can be used to simulate dynamic trajectories and to perform *in-silico* perturbation experiments. We have extensively used discrete-state network modeling to investigate virus infections and have experimentally validated the predictions (36, 37). Here, we extensively expand our method and present single-cell Boolean Omics Network Invariant-Time Analysis (scBONITA) to (a) infer Boolean rules describing signal integration for pathway topologies using scRNA-seq data and (b) use these rules to identify dysregulated pathways and to prioritize genes/proteins for further investigation. scBONITA returns precise modes of dysregulation, captured by node-specific scores that quantify the contribution of each node to the overall dysregulation of a pathway. scBONITA’s capability to perform *in silico* simulation and perturbation of molecular pathways shows that it is a powerful hypothesis-generating tool.

Here, we describe the immune cell subpopulations and subpopulation-specific gene expression programs in a novel scRNA-seq dataset obtained from PLWH with and without atherosclerosis. We use scBONITA to identify dysregulated pathways in individual cell subpopulations, focusing on CD8+ T cells and monocytes. scBONITA highlights dysregulated cell migration and lipid metabolism pathways in several subpopulations and influential genes, such as PI3K and PLC, which have high impacts on signal flow in these networks. Furthermore, we used a publicly available dataset of PBMCs from persons before and after HIV infection (38) to show that cell migration pathways are also dysregulated in the early stages of HIV infection, suggesting modulation of these pathways both by HIV infection and in subsequent AS. We present a novel method (‘attractor analysis’) that uses the models learned by scBONITA to identify pathway-specific signaling states for CD8+ T cells and monocytes. Additional in silico experiments show that scBONITA can learn a context-specific rule set from the vast possible state space for a Boolean network and that scBONITA’s importance score provides novel information about signaling flow. Thus, this work provides insights into the mechanisms of HIV-associated atherosclerosis at the single-cell level by using a novel network modeling algorithm.

## Results

### PBMC subpopulations in AS+ and AS- PLWH

To investigate dysregulated immune signaling in People Living with HIV (PLWH), we recruited a cohort of eight PLWH, four with atherosclerosis (AS+) and four without atherosclerosis (AS-). Participants were matched for known atherosclerosis risk factors (see Methods, Supplementary Note 1, and Supplementary Figure 1). We transcriptionally profiled ∼1200 peripheral blood mononuclear cells (PBMCs) per subject. This data was processed using the Cell Ranger and Seurat pipelines (39) to identify sixteen subpopulations of immune cells (Methods, Figure 1A) annotated using CIBERSORT (40) (Supplementary Figure 2) and cell-lineage specific markers (Supplementary Table 1). This scRNA-seq dataset is referred to in the text as the HIV/AS dataset.

**Figure 1:**
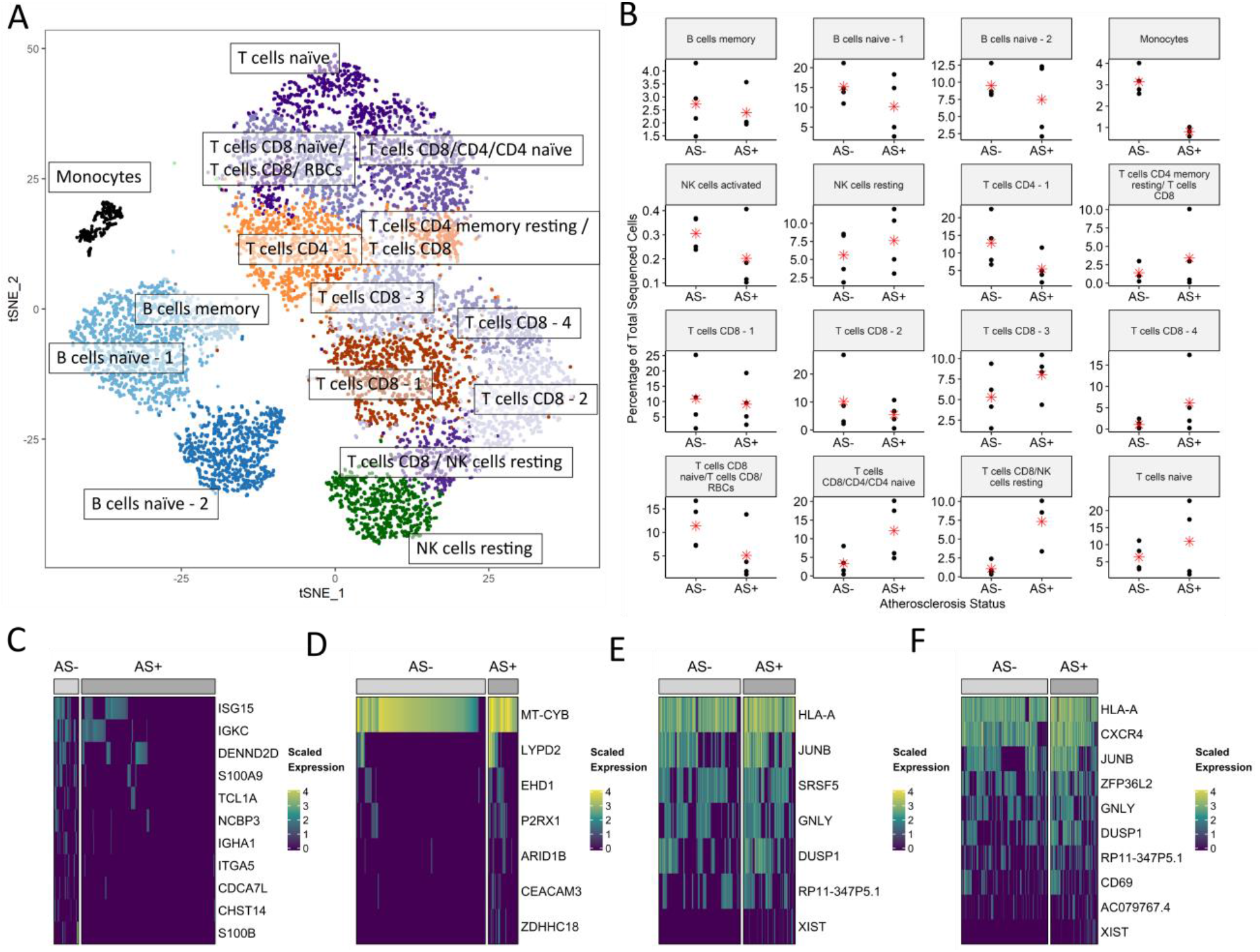
Characterization of PBMC subpopulations in people living with HIV (PLWH) with (AS+) or without atherosclerosis (AS-) (A) t-SNE projection of 16 transcriptionally distinct cell subpopulations, shown in distinct colors. (B) Subpopulation-level differences in the percentage of sequenced cells corresponding to each cell type in panel A from a subject between AS+ and AS- PLWH are identified using a t-test. The mean of each group is represented by a red asterisk. Panels C - F show the expression of genes that are differentially expressed (DE) between cells derived from AS+ and AS- subjects. DE genes were identified using the Wilcoxon test (Bonferroni-adjusted p-value < 0.1, absolute log2 fold change > 0.3.) DE genes in (C) CD8 T cells/NK resting cells, (D) monocytes, (E) naïve B cells referred to as “B cells naïve - 2” in panels A and B, and (F) T cells referred to as “T cells CD8/CD4/CD4 naïve” in panels A and B.

A population of CD8 T cells/NK resting cells was lower in AS- PLWH and a population of CD14+CD16+ monocytes was higher in AS- PLWH (t-test, p < 0.05) (Figure 1B). Cell subpopulation markers were identified using MAST (41) as described in the Methods. The CD14+CD16+ monocytes and CD8+ T cells/NK resting cell markers were enriched for migration-related pathways (Supplementary Table 1). Indeed, these cells are known to migrate into the intima during the formation of atherosclerotic lesions in the vascular wall (42–48).

In addition, we evaluated the proportions of the cell subpopulations identified in scRNA-seq data in a bulk RNA-seq dataset obtained from an independent cohort of matched PLWH with and without AS (49). This dataset was deconvoluted using CIBERSORT (40, 50) to quantify the abundance of the cell subpopulations found in the scRNA-seq dataset (Supplementary Figure 3). We found that most clusters were not significantly different between AS+ and AS- groups, as observed in scRNA-seq data. The subpopulation ‘T cells CD8 NK cells resting’ was more abundant in AS- PLWH (t-test, p < 0.1), while there was no significant difference in the abundance of monocytes. As these differences in the abundance of cell subpopulations were not robustly recapitulated in the independent cohort, here we focus on differentially regulated molecular mechanisms between AS+ and AS- PLWH.

### Atherosclerosis-associated gene-expression across PBMC subpopulations

Differentially Expressed (DE) genes between AS+ and AS- PLWH (Figure 1C-F and Supplementary Table 2) include upregulation of MHC Class I and Class II genes in multiple cell subpopulations. ITGB2, which is upregulated in CD8+ T cells from AS+ PLWH, is involved in leukocyte transendothelial migration (Supplementary Table 2). ACTB (β-actin) was upregulated in naïve B cells and CD8+ T cells from AS+ PLWH (Supplementary Table 2). CXCR4 was upregulated in cells from AS+ PLWH in naïve B cells, CD8+ T cells, and a population of resting NK cells. Both CXCR4 and ACTB modulate dynamic actin cytoskeleton remodeling in transendothelial migration. Gene set ORA of DE genes revealed enrichment of cell migration and mobility functions in cells from AS+ PLWH (Supplementary Table 2).

In AS- PLWH, S100A8/S100A9 are upregulated in three cell subpopulations (Figure 1C-F, Supplementary Table 2). S100B is upregulated in CD8+ T cells populations from AS- subjects (Figure 1C, 1F, Supplementary Table 2). Several ribosomal genes were DE in twelve subpopulations; out of which they were only upregulated in naïve B cells from AS- PLWH and naïve T cells from AS+ PLWH (Supplementary Table 2).

While DE and enrichment analysis indicated some mechanisms of HIV-associated atherosclerosis, an integrated model of how these genes jointly regulate signaling cascades did not emerge. Hence, we developed the network-based pathway analysis algorithm scBONITA (Single-Cell Boolean Omics Network Invariant-Time Analysis) to investigate signal integration and flow.

### scBONITA learns discrete-state models of signaling pathways

scBONITA is a discrete-state modeling approach to develop executable models of dysregulated immune signaling pathways (35) (Figure 2). The scBONITA algorithm requires two inputs: (a) a scRNA-seq dataset, and (b) a prior knowledge network (PKN) (Figure 2A). In these PKNs, genes and their regulatory interactions are represented by nodes and directed edges respectively. scBONITA leverages the principle that observed states of single cells correspond to states of dynamic biological networks to identify regulatory rules (Figure 2B). A genetic algorithm is used to identify a minimum-error rule set that is optimized by a node-wise local-search. This returns a set of discrete-state models for pathways as multiple rule sets explain the training data equally well (Equivalent rule set, ERS). A pathway is described as ‘optimized’ if scBONITA-RD successfully reduced the state space of the possible rules for at least one node in the pathway.

**Figure 2:**
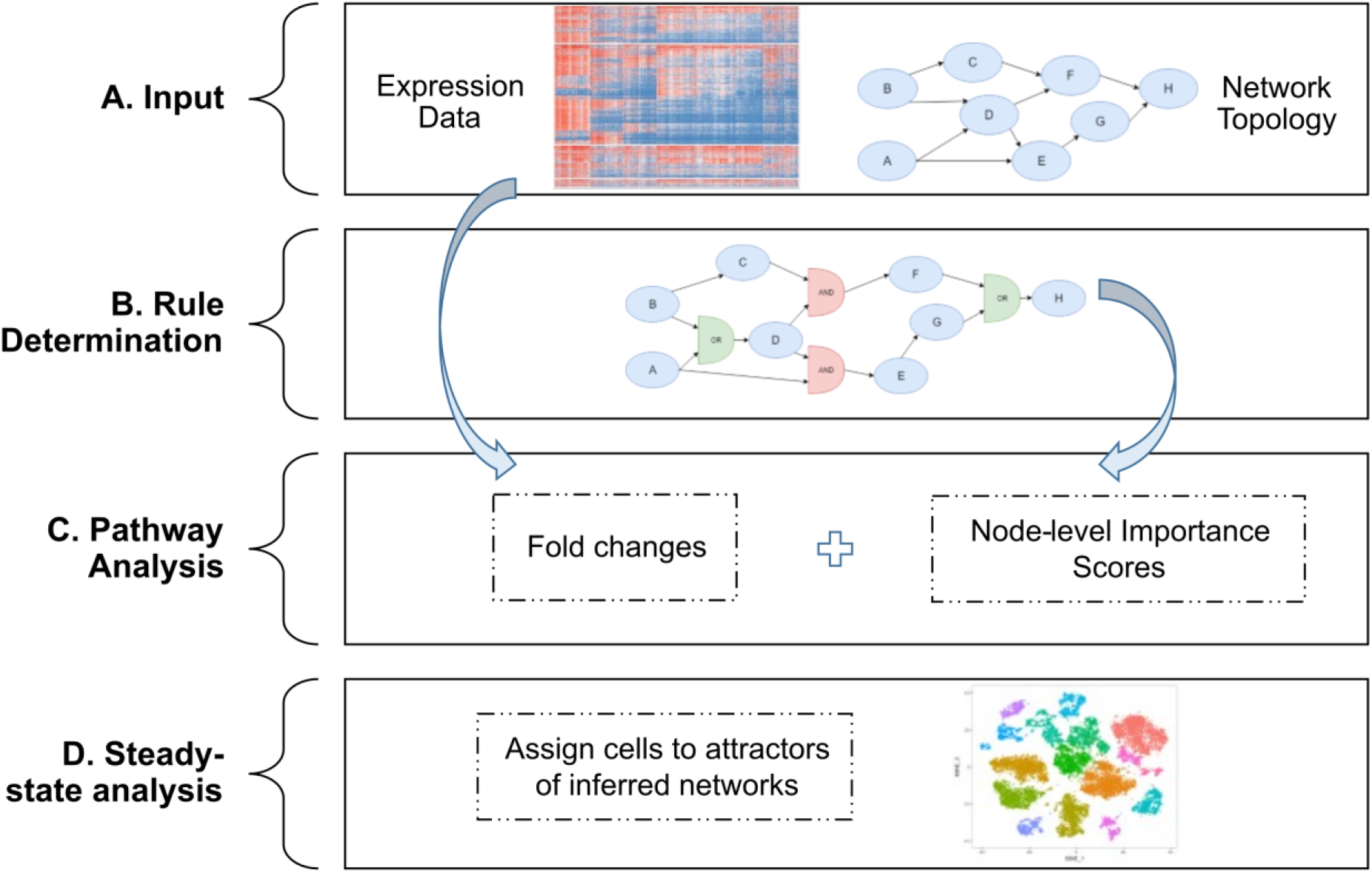
scBONITA pipeline to infer Boolean rules and perform pathway analysis using single cell expression measurements (A) **Input**: a binarized single-cell RNA-seq dataset as a text file, and a prior knowledge network (PKN) describing the activating or inhibitory relationships between genes (B) **Rule determination:** inference of logic rules that describe the regulatory relationships between nodes in the PKN by a global search followed by node-level rule refinement (C) **Pathway analysis:** scBONITA calculates a gene importance score calculated by simulating network perturbations with inferred rules and combines these scores with fold-changes from scRNA-seq to identify differentially regulated pathways in a specified contrast (D) **Steady-state analysis**: scBONITA simulates networks using learned rules to identify steady states which correspond to observed cellular states.

scBONITA models can be simulated to generate dynamic trajectories and *in silico* node perturbations. These models are used to perform node knock-out and knock-in simulations and the difference between network states after these simulations is weighted by the size of the ERS to calculate a node importance score. Thus, node importance score measures the influence of a node over the network and the weight incorporates the uncertainty in rule determination. scBONITA combines these importance scores and comparison-specific fold changes to calculate a pathway modulation score (Methods, Figure 2C).

The simulation trajectories of these discrete-state models fall into steady states known as attractors, which have been hypothesized to correspond to signaling behavior characteristic of specific cell types (51–56). Cells are assigned to the attractor closest to their expression (Figure 2D) to characterize signaling states for a network. In conclusion, scBONITA allows in depth comprehension of signaling pathways by incorporating network topology.

### Dysregulated pathways in T cell populations linked to HIV-associated atherosclerosis

The scBONITA pathway analysis algorithm identifies dysregulated pathways in all subpopulations derived from AS+ and AS- PLWH, providing insights into mechanisms of HIV-associated atherosclerosis (Supplementary Table 3). In CD8+ T cells, these pathways included pro-inflammatory, anti-viral, cell migration and apoptosis pathways (Figure 3A, Supplementary Table 3). all of which were downregulated in AS+ PLWH except the Th17 cell differentiation pathway, which includes genes involved in generic T cell differentiation. All these pathways were identified by scBONITA, but not by enrichr (Supplementary Table 2).

**Figure 3:**
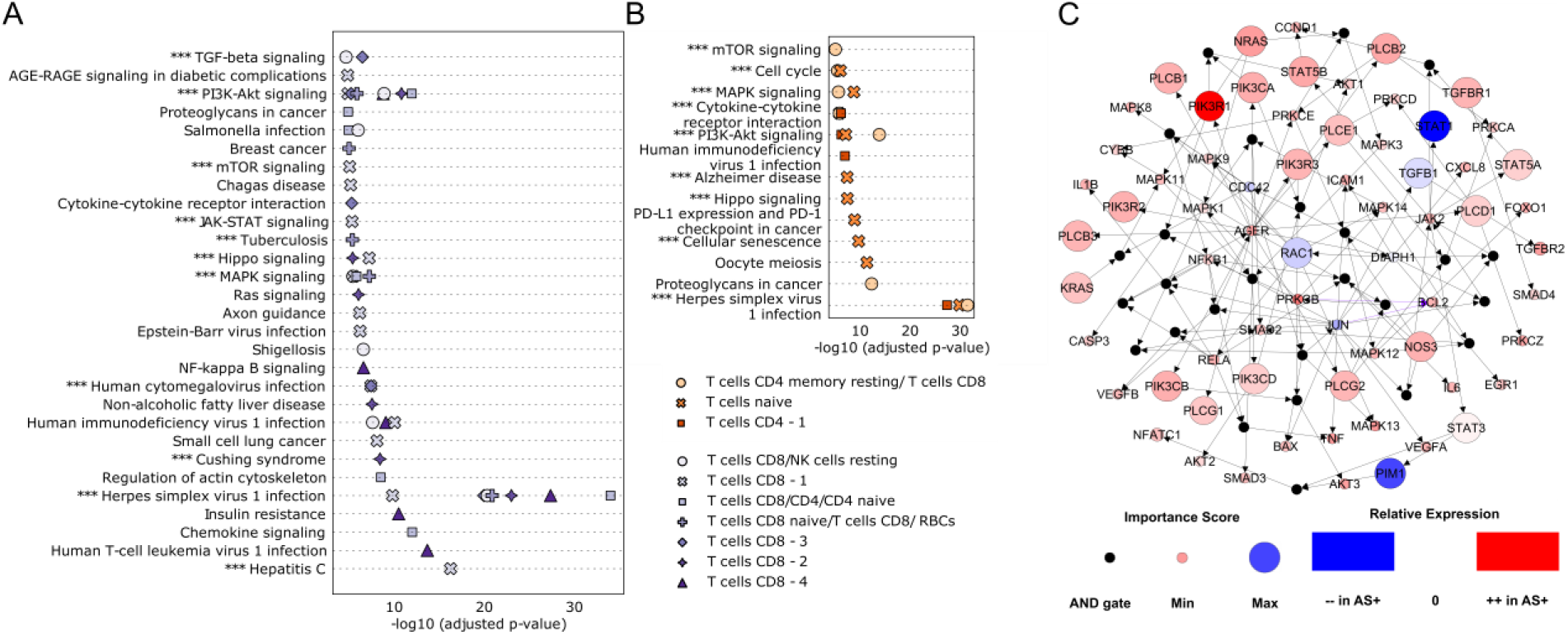
scBONITA identifies dysregulated pathways in T cells derived from AS+ and AS- PLWH. Pathways (y-axis) dysregulated in the AS+ vs AS- contrast in PLWH in clusters of (A) CD8+ T cells and (B) CD4+ T cells and naïve T cells. Clusters are differentiated by point shape, as shown in the legend. Pathways that have Bonferroni-corrected p-value < 0.01 (x-axis) and a reduced ERS (see Methods for details) are shown. Pathways labeled with “***” were also dysregulated between cytotoxic T cells (A) and T cells (B) derived from HIV-subjects and subjects after 1 year of HIV infection (38) (C) Network representation of the AGE-RAGE signaling pathway (Bonferroni-corrected p-value < 0.01) in a cluster of CD8+ T cells referred to as CD8 T cells -1 in Figure 1A. Small black intermediate nodes indicate that the downstream nodes are controlled by an AND function of the upstream nodes. The size of nodes corresponding to genes is proportional to their importance score calculated by scBONITA. Nodes are colored according to the magnitude of their fold change between the HIV+AS+ and HIV+AS- groups. Violet edges indicate inhibition edges and black edges indicate activation edges.

scBONITA identified multiple optimized pathways in CD4+ T cells (subpopulations represented in Figure 1A) as being dysregulated (p_adj_ < 0.01) in AS+ PLWH (Figure 3B, Supplementary Table 3). CD4+ T cells may exert either an atherogenic or atheroprotective phenotype, depending on subset and interactions with antigen presenting cells in the adventitia or plaques (57). Our results suggest that CD4+ and CD8+ T cells from AS+ PLWH play a role in cell adhesion, apoptosis and migration processes involved in atherosclerosis, and these atherogenic processes are mediated by the upregulated PI3K-AKT, mTOR and cytoskeletal signaling pathways.

The AGE-RAGE signaling pathway was further investigated in CD8+ T cells because of its role in dysregulated lipid metabolism (19, 23, 58–63) and to demonstrate the additional information obtained from scBONITA in comparison to ORA. This pathway had the highest pathway modulation score (0.8) (Supplementary Table 3). Most genes in this pathway were upregulated in AS+ PLWH (Figure 3C). scBONITA optimized rules for DIAPH1 (uncertainty score = 0.5, see Methods) (Figure 3C), which has a strong influence over the signal flow through the network due to its high centrality. The combination of scBONITA’s node importance score and fold change between subject groups was used to identify genes whose activity influences signal flow in AS+ PLWH. These genes include: the class 1 PI3K genes, the P13K regulator PI3KR1, and PLC genes. All of them are highly expressed in AS+ PLWH (Figure 3C). PI3K activates lipid metabolism, macrophage autophagy, phenotypic transition, and the expression of adhesion molecules (reviewed in (64)). In this manner, scBONITA identified dysregulated pathways and genes associated with atherosclerosis-linked migration of T cells derived from PLWH.

### Dysregulated pathways in monocytes linked to HIV-associated atherosclerosis

In monocytes, scBONITA identified several dysregulated pathways (Figure 4A, Supplementary Table 3) which are known to be involved in the pro-inflammatory behavior of pro-atherogenic monocytes (43, 65–74). Of these pathways, only cAMP signaling and endocrine resistance are overall upregulated in cells from AS+ PLWH, suggesting that monocytes from AS- are pro-atherogenic.

**Figure 4:**
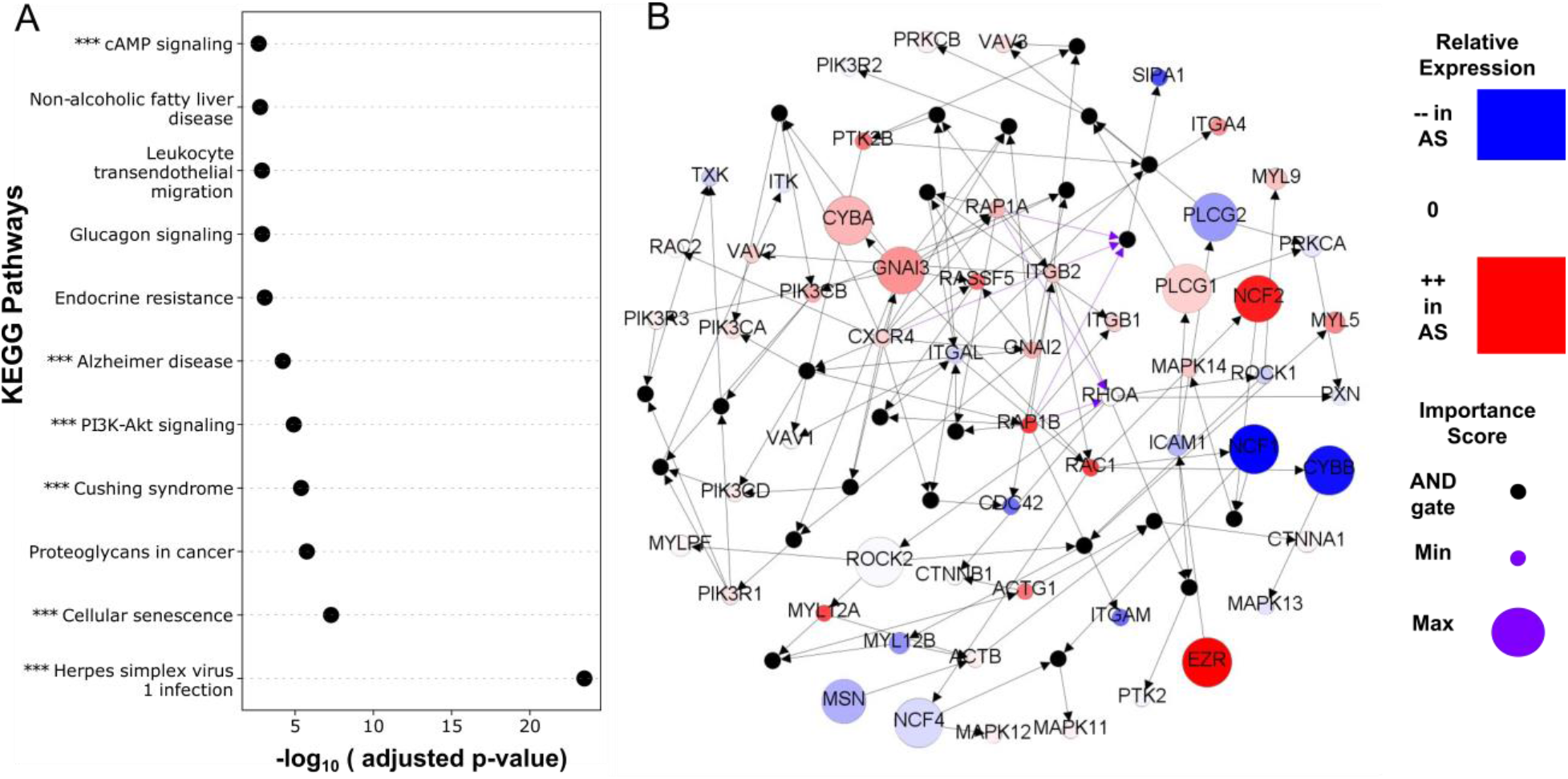
scBONITA identifies dysregulated pathways in monocytes derived from AS+ and AS- PLWH: (A) Pathways (y-axis) dysregulated in the AS+ vs AS- contrast in monocytes derived from PLWH. Only pathways that have Bonferroni-corrected p-value < 0.01 (x-axis) and which have a reduced ERS (see Methods for details) are shown. Pathways labeled with “***” were also significantly dysregulated in monocytes after one year of HIV infection (38) (B) Network representation of the leukocyte transendothelial migration pathway. Small black intermediate nodes indicate that the downstream nodes are controlled by an AND function of the upstream nodes. The size of nodes corresponding to genes is proportional to their importance score as calculated by scBONITA. Nodes are colored according to the magnitude of their fold change between the HIV+AS+ and HIV+AS- groups. Violet edges indicate inhibition edges and black edges indicate activation edges.

The leukocyte transendothelial migration pathway was further investigated as monocyte migration outside the vascular compartment plays a crucial role in the inflammatory cascade that leads to an atherosclerotic phenotype (65, 67, 75). The pathway modulation score of 0.45 was the third highest amongst tested pathways (Supplementary Table 3) (Figure 4B). scBONITA optimizes regulatory rules for the influential RHOA gene (uncertainty factor=0.13). High importance scores were assigned to the NCF genes, PLCG genes, MSN, ROCK, CYBA and CYBB. NCF genes are involved in superoxide production and are a positive regulator of P13K signaling (76, 77). The upstream regulator of PLCG1, MSN, is involved in cytoskeletal remodeling during leukocyte migration (78). ROCK2 shows a small change across AS groups, which may be driven by feedback regulation, but has a stronger role in regulating signal flow, as shown by its high importance score. The high importance of the G protein GNAI3, downstream of CXCR4 indicates a role for CXCR4-mediated activation of this pathway. GNAI3 is also present in two other significantly dysregulated networks in monocytes – Cushing Syndrome and cAMP signaling – but does not have a high importance score in those networks, indicating that it is critical in regulating signal coming from CXCR4 only in the leukocyte transendothelial migration network. The downstream effectors of these genes have higher fold changes, possibly due to feedback regulation. These effectors include ACTG1 and EZR, involved in cytoskeletal remodeling (78–80), and ITGA4, ITGB1, and ITGB2, involved in cell adhesion. scBONITA thus identifies several genes and pathways associated with dysregulated transendothelial migration in the context of HIV-associated atherosclerosis.

### Pathways dysregulated by HIV infection and in HIV-associated atherosclerosis

To identify the biological mechanisms modulated by HIV infection that may also contribute to the elevated risk of AS in PLWH, we used a publicly available scRNA-seq dataset of PBMCs from persons before and during acute HIV infection (38). Kazer et al (38) identified gene expression programs in activated T cells, monocytes, and NK cells during HIV infection. We used this dataset with scBONITA to infer Boolean rules for KEGG networks and perform pathway analysis. The results were compared to the pathways dysregulated in HIV-associated atherosclerosis in the above-described HIV/AS dataset.

scBONITA identified 10 optimized pathways dysregulated after 1 year of HIV infection in cytotoxic T cells (38) (Figure 3A-B & 5A, Supplementary Figure 6, Supplementary Tables 5 & 6). Of these pathways, five signaling pathways were dysregulated in AS+ PLWH. Similarly, 5 out of 19 optimized pathways were dysregulated in monocytes upon HIV infection and in AS+ individuals (Figures 4A & 6A, Supplementary Figure 6, Supplementary Table 6). 81 and 41 genes from these overlapping pathways were upregulated both upon HIV infection and in AS+ PLWH in the cytotoxic T cell populations and monocyte population respectively (Supplementary Figure 7, statistical significance was not tested as scBONITA does not depend on DE genes to perform pathway analysis). The genes upregulated after HIV infection and in AS+ PLWH in the CD8+ T cell subpopulation were involved in viral response pathways (Supplementary Figure 7). Similarly, the genes upregulated after HIV infection and in AS+ PLWH in the monocyte subpopulation are involved in cell migration related pathways (Supplementary Figure 7). This suggests that the modulation of cell migration and inflammation processes upon HIV infection progresses over time, increasing risk of AS in PLWH.

**Figure 5:**
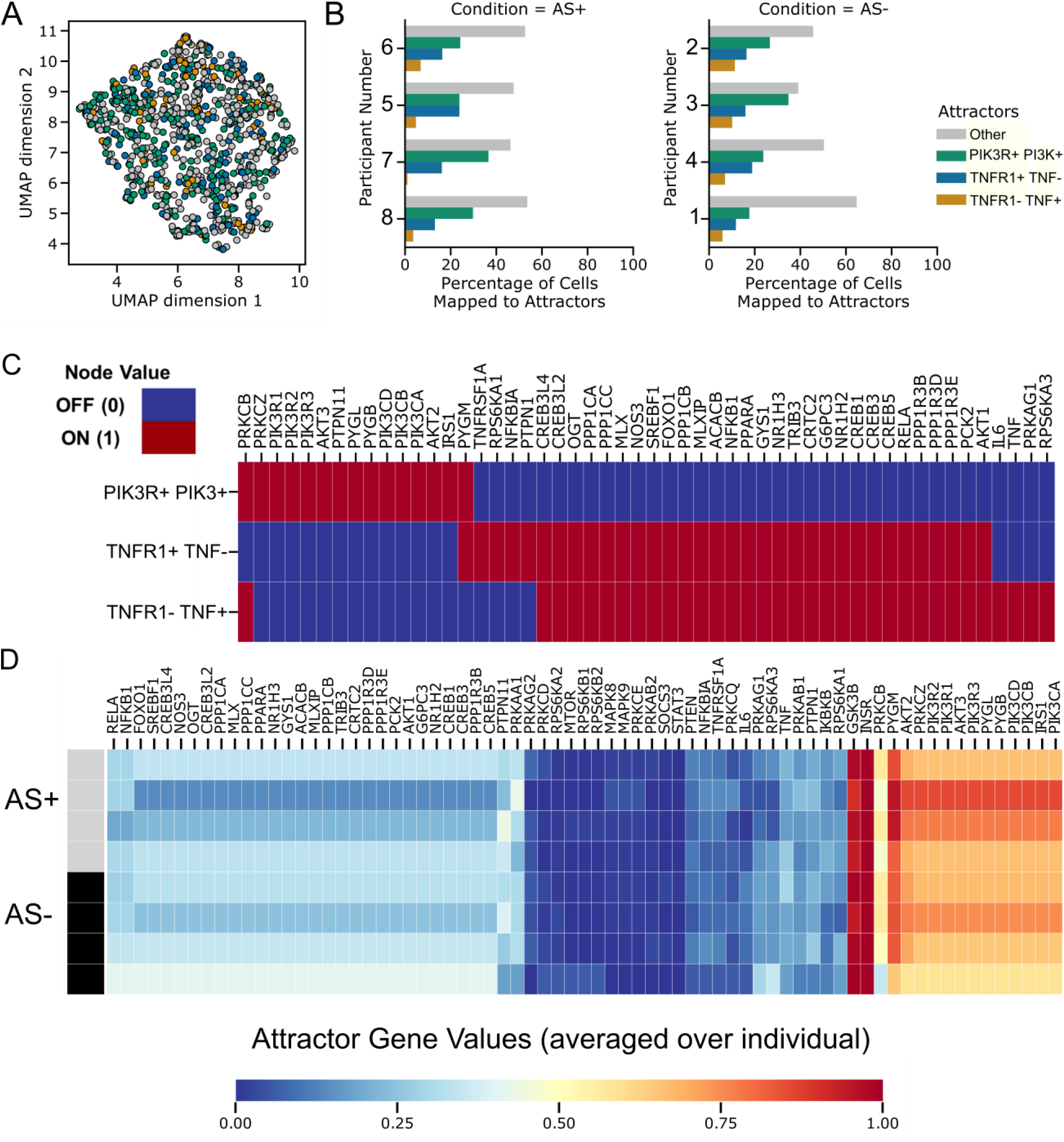
CD8+ T cell states with respect to the insulin resistance pathway identified by attractor analysis with scBONITA. (A) UMAP representation of a cluster of CD8+ T cells (CD8+ T cells – 1 in Figure 1A) colored by the attractor to which they are assigned, based on their similarity. The three dominant states (PI3KR+ PI3K+, TNFR1+TNF-and TNFR1-TNF+ attractors) are represented by green, blue and orange. All other attractors are collectively labeled in grey. (B) Percentages of CD8+ T cells derived from each subject, mapping to the three dominant and all other attractors. (C) Gene activity (ON-red, OFF-light blue) in the three dominant attractors. Only genes that are different between these states are shown. (D) Attractor gene values ranging from 0 (blue) to 1 (red) averaged for each individual subject. The top bar indicates AS+ (grey) and AS- (black) subjects.

**Figure 6:**
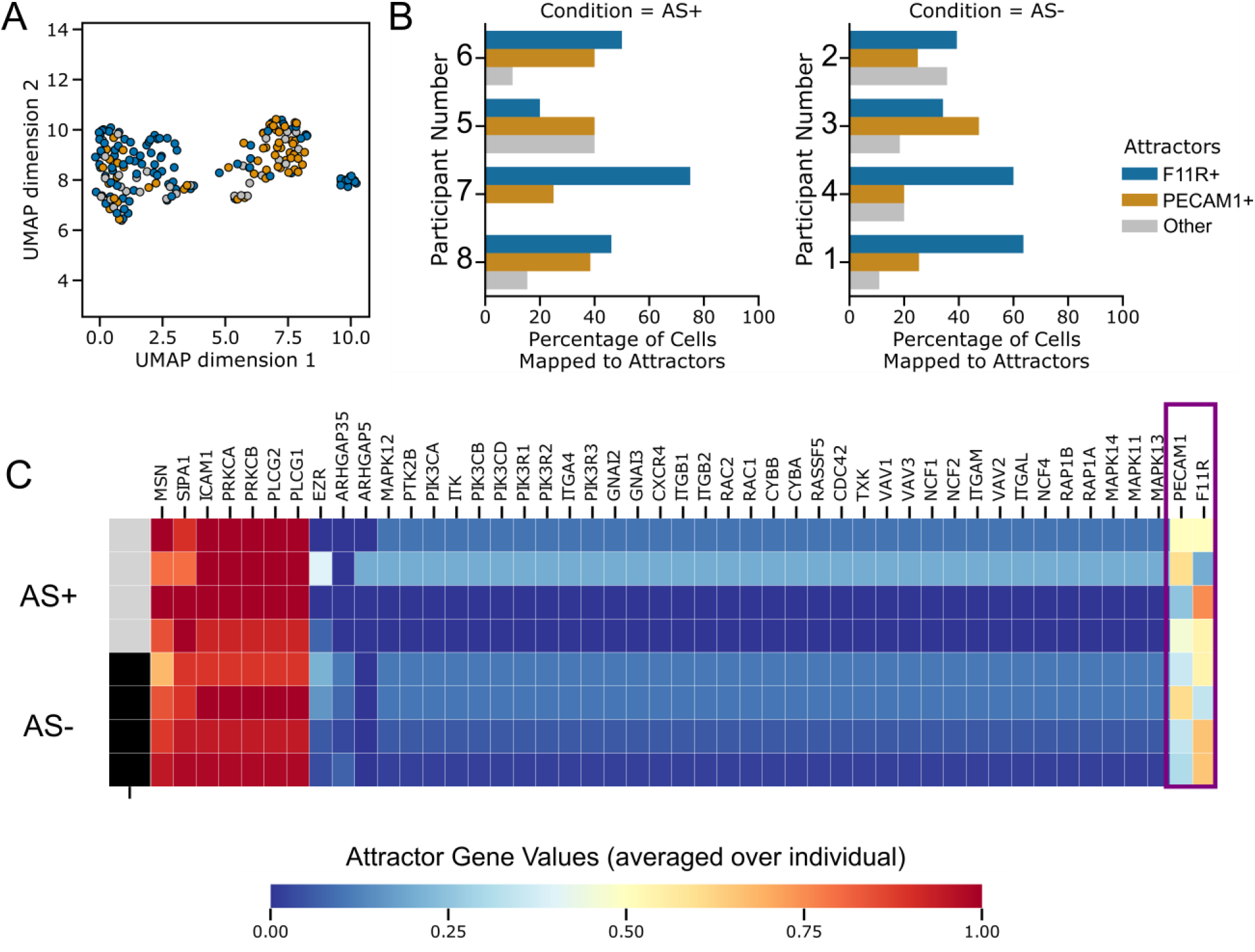
Monocyte states with respect to the leukocyte transendothelial migration pathway identified by attractor analysis with scBONITA. (A) UMAP representation of the cluster of monocytes colored by the attractor to which they are assigned, based on their similarity. The two dominant modes (F11R+ and PECAM+ attractors) are represented by blue and orange. All other attractors are collectively labeled in grey. (B) Percentages of monocytes derived from each subject, mapping to the two dominant attractors and all other attractors for the leukocyte transendothelial migration pathway. (C) Attractor gene values for the leukocyte transendothelial migration pathway trained on monocytes, ranging from 0 (blue) to 1 (red), averaged for each individual subjects. The top bar indicates AS+ (grey) and AS- (black) subjects. The genes that differ between the two dominant attractors F11R+ and PECAM+ are highlighted by a violet box.

### Pathway-specific cellular signaling states associated with atherosclerosis

We used the network models learned by scBONITA to identify attractors for the pathways dysregulated between AS+ and AS- PLWH in CD8+ T cells discussed above (Cluster CD8 T cells -1 in Figure 1A) and evaluated cellular states across subjects and disease groups. The simplest rules, which have the smallest number of “AND” terms, were chosen to simulate the network and identify attractors (Methods). The insulin resistance pathway, which is downstream of the AGE-RAGE and PI3K-AKT signaling pathways, was particularly interesting in CD8+ T cells (Cluster CD8 T cells -1 in Figure 1A). 72 signaling states were identified by network simulation, of which 3 dominant signaling states mapped to 16.5%, 7.5% and 27.4% of cells respectively (Figure 5A-B). There is an association between attractors and subjects (chi-square test, p-value < 0.05) but no association between attractors and atherosclerosis status (chi-square test, p-value > 0.05). Additionally, we also performed attractor analysis for other cell clusters. Among those, interestingly, assigned chemokine signaling attractors of CD4+ T cells were subject-specific (chi-square test, p < 0.01). Similarly, we observed subject-specific attractors of the PI3K-AKT signaling pathway for the T cells CD8/CD4/CD4 naive cluster (chi-square test, p < 0.01).

The CD8+ T cell states for the insulin resistance pathway were characterized by differences in several key genes (Figure 5C), including PI3K genes and the PI3K regulators that were identified as being influential in the AGE-RAGE signaling pathway (Figure 3B). In addition, the two less abundant attractors differed in the activity of the key TNFR and TNF genes. Hence, these attractors are referred to as the PIK3R+ PI3K+, TNFR+TNF- and TNFR-TNF+ attractors. The activity of PI3K and AKT genes was higher in the most common signaling state (PI3KR+ PI3K+ attractor). However, the activity of the downstream targets of AKT, such as the CREB genes, NFKB1, FOXO1, were lower in the PI3KR+ PI3K+ attractor. TNF, which is produced at a low level by some subsets of T cells, also mediates a range of pro-inflammatory processes in vascular endothelial cells (81, 82). TNF-TNFR1 signaling mediates an apoptotic process mediated by TRADD and FADD (83). TNFR1 activates PI3K signaling in regulatory T cells (84). Thus, our attractor analysis reveals different T CD8+ cell states. This analysis also indicates differences in TNF production and response in the cells that are assigned to these cell states and suggests the existence of distinct modes of operation across subjects (Figure 5D).

Attractor analysis of the leukocyte transendothelial migration pathway in monocytes revealed 9 attractors that mapped to cells in the dataset. The two dominant signaling modes differed in the activity of the PECAM1 and F11R genes respectively (Figure 6A-C). The attractors are referred to as the PECAM+ and F11R+ attractors and mapped to 51.04% and 30.73 % of cells respectively. F11R is required for platelet adhesion to vascular endothelial cells (85), which occurs prior to infiltration of monocytes into the endothelium and eventual plaque formation (86, 87). PECAM1 has widespread effects on vascular biology and atherosclerosis in particular (88–90). These subject specific attractors (chi-square test, p-value < 0.05, Figure 6B) were not associated with atherosclerosis status. Similarly, the attractor activity of individual genes was not associated with atherosclerosis status (two-sided t-test, p-value > 0.05). Thus, scBONITA identifies two cellular states of monocytes driven by molecular signaling. These states are associated with monocyte migration into the vasculature and reflect inter-subject variability.

### Evaluation of scBONITA performance in silico

The BONITA algorithm has already been rigorously validated in our previous study (35). Specifically, comparison with other pathway analysis tools has been performed. Here we evaluate scRNA-seq specific components of the algorithm. To show that scBONITA rule determination is robust to training set size, we varied the size of the training data provided to scBONITA. The number of cells in the largest cluster of cells (Naïve B cells -1) were varied by random selection from 1% of cells from that cluster to 200% by adding cells from neighboring clusters. The reduced size of the ERS for nodes with in-degree 3 (i.e., the most complex case considered by scBONITA) is a metric for certainty in rule inference by scBONITA (Figure 7A). While there was a significant decline in performance when the data was downsampled to 1% of the original cluster, there was no significant increase in effect once 50% of the cells were used, or when the training dataset was augmented. This indicates that scBONITA is robust to heterogeneity in the training data set.

**Figure 7:**
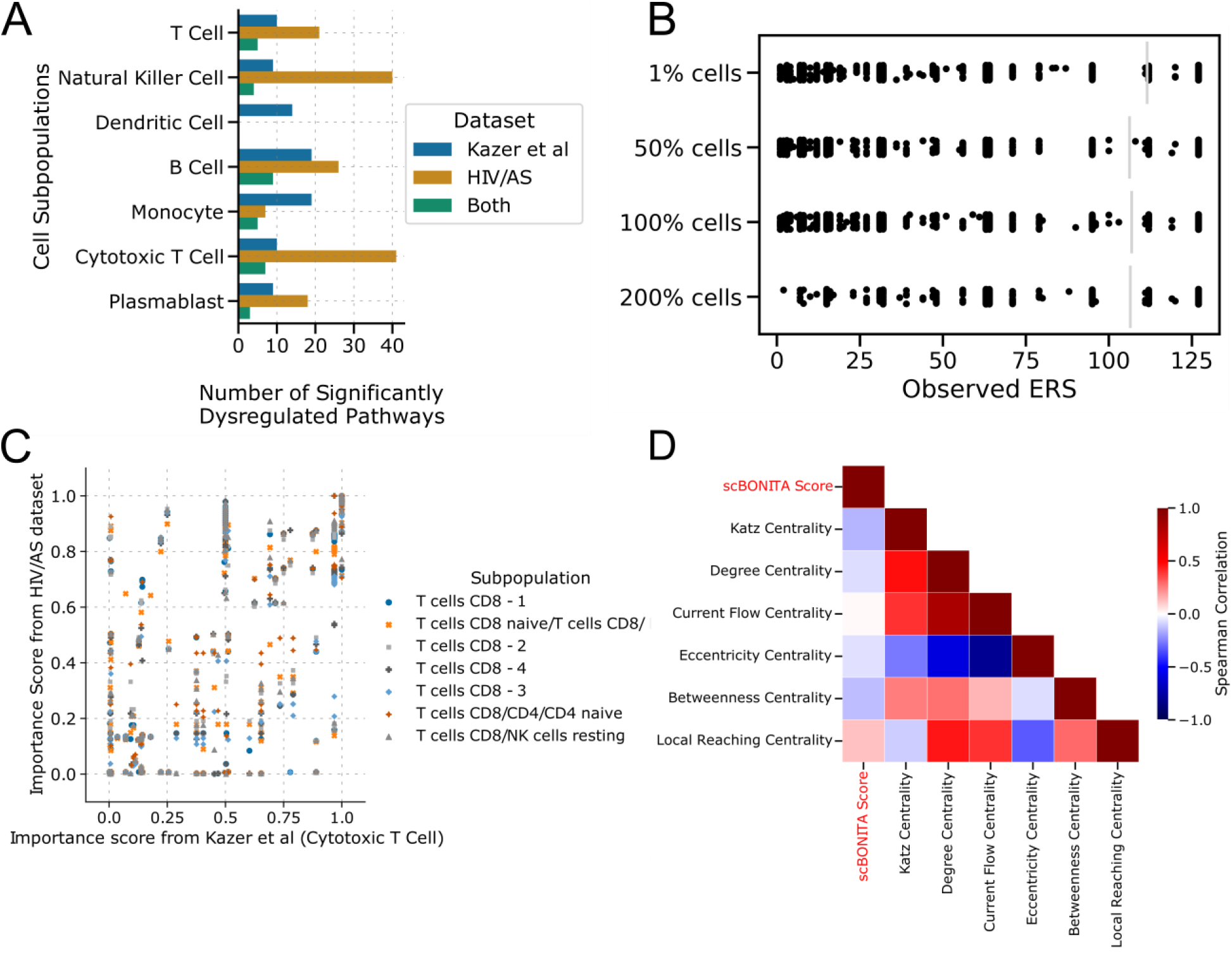
Performance of scBONITA rule determination. (A) The number of pathways identified as significantly dysregulated (Bonferroni-adjusted p value < 0.05) one year upon HIV infection (38), between AS+ and AS- PLWH, and the intersections between these sets. Subpopulations from the two datasets were matched as shown in Supplementary Table 5. (B) Effects of number of cells on the ERS size evaluated by downsampling and augmentation using the largest cluster (“B cells naïve – 1”) from the HIV/AS dataset (C) Relation between importance scores in 130 KEGG networks evaluated using CD8+ T cells from two comparisons - AS+ and AS- PLWH and from persons before and one year after HIV infection (38). (D) Spearman correlations (p < 0.01 for all comparisons) between scBONITA’s node importance score (labeled as ‘scBONITA score’) and 6 measures of node centrality (along x and y axis). Correlation coefficients are depicted by colors ranging from blue (-1) to red (+1).

The node importance scores calculated by scBONITA for KEGG networks trained on the HIV/AS dataset as described above are not correlated to six centrality measures (Figure 7D). We found that the node importance scores between comparable cell subpopulations in the Kazer et al and HIV/AS datasets investigated here were correlated, as shown by a representative comparison between cytotoxic T cells from the Kazer et al dataset and the subpopulations of CD8+ T cells from the HIV/AS dataset (Figure 7C, 0.71 < Pearson Correlation Coefficient < 0.91, p < 0.01). Similarly, the node importance scores for the populations of monocytes were highly correlated (Pearson correlation coefficient = 0.78, Supplementary Table 4, Supplementary Figure 4). The correlations were lower for other pairs of subpopulations (Supplementary Table 4, Supplementary Figure 4). This indicates that scBONITA learns some characteristic features of network topologies, but node importance scores are still assigned in a context-dependent manner.

## Discussion

Among people living with HIV, widespread use of cART has significantly reduced overall mortality. However, the earlier and increased incidence of cardiovascular diseases remains the major cause of mortality in an aging HIV+ population for multiple intersecting reasons (4, 9–12). We and others have attempted to identify the immune signaling mechanisms that lead to this increased incidence of atherosclerosis (49). However, to the best of our knowledge no study has systematically investigated cell-type specific signaling dysregulations at single-cell level. To this end, we sequenced, for the first time, PBMCs from four AS+ and four AS- PLWH. The cohort was closely matched for known AS risk factors. These risk factors will be harder to match exactly in a larger cohort. Although a small sample size in some contexts can limit the power of the study, previous scRNA-seq studies have provided insight into both changes in PBMC composition and signaling with cohort sizes similar to this study in the context of HIV, because of the depth of information on molecular mechanisms that can be obtained from scRNA-seq experiments (38, 91–94). To increase statistical power, we evaluated the proportion of the subpopulations identified from scRNA-seq in a bulk RNA-seq dataset from an independent cohort of PLWH with and without AS (49) (Supplementary Figure 3). Enrollment of more subjects in the future will help further elucidate the signaling dysregulations leading to HIV-associated atherosclerosis. We found that in accordance with previous studies (19, 58–62, 95, 96), a population of CD8+ T cells was increased in PBMCs from AS+ PLWH. Contrary to our expectations (20, 66, 67, 72, 74, 97–100), a population of monocytes was decreased in PBMCs from AS+ PLWH. Here we focus on cell-type specific molecular changes associated with atherosclerosis.

We observed that differentially expressed genes between AS+ and AS- were involved in cell migration; however relevant pathways were not identified using over-representation analysis (ORA). This demonstrated the inefficacy of conventional ORA methods in identifying coherent molecular processes. For example, CXCR4, a DE gene, leads to the activation of phosphatidylinositol-3-OH kinases (PI3K), which in turn activates the serine-threonine kinase AKT via PIP3 (101). PI3K/AKT signaling leads to processes involved in plaque formation, such as cell migration, intracellular lipid accumulation, and smooth muscle cell proliferation (64). None of these processes were enriched in the DE genes. This may be because of the technical limitations of scRNA-seq leading to nonspecific distortions of expression and identification of fewer differences across conditions (102, 103). Even DE analysis methods that are sensitive to the known characteristic distributions of scRNA-seq data are prone to false discoveries (104). These methods failed to provide insights into how disparate genes involved in different pathways regulate cellular states. To mechanistically characterize signaling dysregulations in HIV-associated atherosclerosis, we developed the scBONITA algorithms for regulatory rule inference, network simulation, pathway analysis, and attractor/steady-state analysis. As a gene is evaluated in the context of its complete signaling pathway, our approach minimizes the impact of the caveats in scRNA-seq technology described in (102, 103).

scBONITA learns condition-specific logic models using scRNA-seq data in conjunction with published prior knowledge networks (PKNs). This study builds on our published BONITA method (35) that inferred logic rules from bulk RNAseq data. scBONITA exploits the bimodal nature of scRNA-seq data (105, 106) and the cell-level resolution of expression to successfully learn regulatory rules and identify attractors for PKNs. These rules can be used to perturb and simulate pathways *in silico*. Unlike other algorithms to infer logic rules and reconstruct gene-regulatory networks on a small subset of genes from scRNA-seq data (107–109), scBONITA does not pre-select genes. In addition, scBONITA is not dependent on time-series data, a significant advantage since time-series data is rarely available in human studies. In lieu of using time-series data, scBONITA hypothesizes that scRNA-seq data represents samples in the state space of a dynamic Boolean network. In addition, the use of known network topologies reduces the uncertainty of inferred rules. Indeed, we show that scBONITA can successfully restrict the state space of possible rules. scBONITA combines expression information with the importance score to create a unique metric of pathway dysregulation. scBONITA node scores can be directly translated into empirical perturbation studies.

In CD8+ T cells we identified AGE-RAGE signaling, which elicits activation of multiple intracellular signaling pathways such as cell proliferation and apoptosis pathways (Figure 3A) (110–118). The PI3K family of genes, which had the highest importance score and were upregulated in AS+ PLWH, promote intracellular lipid deposition leading to the formation of foam cells and atherosclerotic plaques and can reduce the expression of lipid transporters and reduce the efflux of intracellular cholesterol depending on upstream signals (64). PLC, which also has a high importance score, promotes leukocyte adhesion, VEC apoptosis and plaque development induced by oxidized low-density lipids (78, 119).

In monocytes, dysregulation of lipid-metabolism pathways such as cAMP signaling and leukocyte transendothelial migration suggest the infiltration of monocytes into the intima during the formation of atherosclerotic lesions and progression of atherosclerosis (42–47). scBONITA identified genes critical to the atherosclerotic process in the leukocyte transendothelial migration pathway. ROCK activation by atherogenic stimuli such as oxidized LDLs leads to pathophysiological changes include endothelial dysfunction and vascular remodeling (120, 121). ROCK inhibitors such as statins attenuate atherosclerosis by inhibiting chemotaxis of macrophages and their transformation into foam cells (122). The dysregulation of glucose metabolism pathways in AS+ PLWH can induce expression of adhesion molecules by VECs and increased monocyte transendothelial migration (64, 123). Thus, here we find migratory and lipid-metabolism pathways and key genes driving atherosclerosis in PLWH.

PI3K-AKT signaling is dysregulated in all cell subpopulations (Figures 4 – 5, Supplementary Table 3). The activation and effector mechanisms of this cascade vary by cell type, as shown by the different dysregulation of linked signaling pathways in different cell types. For example, in two populations of naïve B cells derived from AS+ PLWH, the apelin signaling pathway was upregulated in one and the adipocytokine signaling pathway was downregulated in the other. The cardioprotective effect of apelin is modulated by (amongst other routes) the PI3K-AKT signaling and MAPK signaling pathways (80–82), which are also dysregulated in these naïve B cells. Apelin is also shown to be upregulated in human atherosclerotic coronary arteries and colocalized with markers for macrophages (124, 125).

We find that HIV infection dysregulates several pathways that are further impacted in PLWH with AS (Figure 3A & 4A, Supplementary Figure 6, and Supplementary Table 6). These pathways are suggestive of changes occurring in cell migration of cytotoxic T cells and monocytes (126–129). The overlap also suggests that cAMP and PI3K-AKT signaling is affected during the course of HIV infection.

To map cells to distinct signaling modes of the pathways described above, we developed scBONITA’s attractor analysis functionalities. Attractors are regions in the state space of a dynamic system towards which simulation trajectories are “pulled” and are characteristics of a specific network with a specific set of regulatory rules. These attractors may correspond to observable cell states, or hallmarks of specific phenotypes such as cell type differentiation, disease state, or drug treatment (51–56). These studies show that even simple dynamic models capture rich and nuanced cell behaviors. scRNA-seq allows the study of these dynamic landscapes and their attractors at an unprecedented resolution (56, 130, 131). Attractor analysis with scBONITA allows users to characterize cells based on the dynamic properties of signaling networks, which dictate phenotype. scBONITA identifies these attractors and their master regulators that control the changes between these cell states, providing complex insights into cellular processes.

The importance of cell migration and lipid signaling in the development of HIV-associated atherosclerosis was underscored by attractor analysis in CD8+ T cells and monocytes. In most cases, only one dominant signaling state existed in the cell subpopulation. However, the three dominant signaling modes of the insulin resistance pathway in CD8+ T cells differed in the activity of PI3K and AKT genes and their downstream effectors, such as CREB and FOXO1. This suggests the existence of two distinct modes for this signaling pathway corresponding to a proliferative cell state (activation of PI3K and AKT) and a senescent cell state (transcription of CREB- and FOXO1-controlled genes) (132–134). The insulin signaling pathway exerts immunomodulatory effects on T cells (135). Similarly, we identified two dominant signaling modes (PECAM+ and F11R+) for the leukocyte transendothelial migration pathway in monocytes, suggesting variation in the cell states with respect to this pathway in PLWH. Dysregulation in insulin signaling, which is a risk factor for CVD promotes PECAM1-mediated migration of monocytes into the endothelium (136, 137). PECAM1’s loss contributes to atherosclerosis (90). PECAM1+ cells may contribute to the suppression of inflammatory processes driving atherosclerosis. Thus, our analysis connects molecular processes to cellular states, unlike conventional DE and ORA analyses.

Although the BONITA algorithm has been rigorously validated in our prior publication (138) here we wanted to evaluate the scRNA-seq specific parts of the algorithm. We demonstrated that scBONITA can identify characteristic structural properties of networks and use this in conjunction with expression information to identify dysregulated pathways in a specified condition (Figure 7). Moreover, scBONITA is minimally affected by the heterogeneity of training data and can narrow down the vast state space for a Boolean network (Figure 7A). We expect that scBONITA inference will improve when pure cell populations are sequenced. While scBONITA is not strictly dependent on the clustering method used to classify scRNA-seq data into subpopulations, we used pre-classified subpopulations to reduce variability and to improve the specificity of scBONITA-PA. Additionally, scBONITA-RD requires a longer runtime (<12 hours in our tests) and more powerful computational capabilities than a typical processing and clustering analysis pipeline run on datasets of typical size. These resources are usually available to academic users on computing clusters. In conclusion, we present a novel dynamic network modeling method that yields mechanistic insights into the cellular and immunological processes involved in HIV-associated atherosclerosis.

## Materials and Methods

### Participant cohort summary, sample collection, and storage

Eight men living with HIV and >= 50 years of age on stable combined antiretroviral therapy (cART) for at least 1 year and with viral load <= 50 copies/mL were recruited. All methods were carried out in accordance with University of Rochester guidelines and regulations, and all experimental and study protocols were approved by the University of Rochester Institutional Review Board (#RSRB00063845). Informed consent was obtained from all subjects. Individuals were classified as having atherosclerosis (AS+) if they had plaques on the carotid arteries on ultrasound imaging. Four of the eight subjects were assigned as AS+ and had plaques in both right and left carotid arteries. AS- subjects were aged between 47 and 57 and AS+ subjects were aged between 51 and 66. AS+ subjects had mean serum cholesterol of 161.5 mg/dl (σ = 40.9) and mean serum high-density lipid HDL of 54.7 mg/dl (σ = 16.3). AS- subjects had mean serum cholesterol of 167.7 mg/dl (σ =57.2) and mean serum high-density lipid HDL of 51 mg/dl (σ = 7.7). AS- subjects and AS+ subjects had a mean CD4+ T cell count of 518.5 cells/µl (σ =347.8 cells/µl) and 838.7 cells/µl (σ = 514.5 cells/µl) respectively. De-identified subject information is available in Supplementary Note 1 and Supplementary Figure 1. Subjects were matched for lipid profiles, hypertension status, smoking status, CD4+ T cell counts, and age. In addition, all subjects were treated with cART for at least one year. 30 mls of blood per study participant was collected in ACD vacutainers and was processed within 2 - 3 hours of collection. Peripheral Blood Mononuclear Cells (PBMCs) were isolated using Ficoll density gradient centrifugation. 5 million PBMCs were preserved using RNAlater (Thermo Fisher) and were used for scRNA-seq.

### Single-cell RNA sequencing and data processing

Frozen vials containing cells in RNAlater were thawed quickly in a 37-degree water bath. Cell suspension was transferred to a 15ml conical tube. 10 ml PBS/2% FBS was slowly added. Samples were centrifuged at 1600rpm for 6 min. Washes were repeated for an additional 2 times for a total of 3 washes. Using the MACS Miltenyi Biotec Dead Cell removal kit (PN130-090-101), dead cells were removed using manufacturer’s recommendations. Cells were counted and cellular suspensions were loaded on a Chromium Single-Cell Instrument (10x Genomics, Pleasanton, CA, USA) to generate single-cell Gel Bead-in-Emulsions (GEMs). ScRNA-seq libraries were prepared using Chromium Single-Cell 3’ Library & Gel Bead Kit (10x Genomics). The beads were dissolved, and cells were lysed per manufacturer’s recommendations. GEM reverse transcription (GEM-RT) was performed to produce a barcoded, full-length cDNA from poly-adenylated mRNA. After incubation, GEMs were broken, and the pooled post-GEM-RT reaction mixtures were recovered, and cDNA was purified with silane magnetic beads (DynaBeads MyOne Silane Beads, PN37002D, ThermoFisher Scientific). The entire purified post GEM-RT product was amplified by PCR. This amplification reaction generated sufficient material to construct a 3’ cDNA library. Enzymatic fragmentation and size selection was used to optimize the cDNA amplicon size and indexed sequencing libraries were constructed by End Repair, A-tailing, Adaptor Ligation, and PCR. Final libraries contain the P5 and P7 priming sites used in Illumina bridge amplification. Sequence data was generated using Illumina’s NovaSeq 6000. Approximately 2000 cells were sequenced from each subject. Cell Ranger (version 2.1.1; 10x Genomics) was used for demultiplexing and alignment with default parameters. Reads were aligned to the human reference genome GRCh38 (Ensembl 93). The Seurat R package (version 2.3.4) (39) was used to further process the gene counts obtained from the CellRanger pipeline. Cells that express < 200 genes, > 2500 genes, or > 5% mitochondrial genes were filtered out. Genes expressed in < 3 cells were filtered out. Gene counts were per-cell normalized and log_2_-transformed. These preliminary filtering and selection procedures yielded a set of 9368 sequenced cells, approximately equally distributed between subjects (and hence conditions), and 14017 genes. Note that sample collection, processing and sequencing were performed in one batch, leading to extremely high-quality data where no subject specific patterns were observed.

### Classification into subpopulations using modularity-optimized Louvain community detection, and cluster labeling

Cells were classified into subpopulations using modularity optimized community detection, implemented in the Seurat R package (39). 664 highly variable genes were used to identify 10 principal components that explained the majority of variance in the data. These principal components were used to cluster the data. Clustering yielded 16 subpopulations. Cluster markers were identified using MAST (105). As suggested in (139), CIBERSORT (40) was used to “deconvolute” the average gene expression of each cluster into the constituent canonical cell types. A reference expression set of 22 immune cell types and 547 genes was used (40). Over-representation analysis was performed using the implementation of the hypergeometric test in the R package clusterprofiler (version 3.12.0) with Kyoto Encyclopedia of Genes and Genomes (KEGG) gene sets downloaded from MSigDb (140–142). Gene sets were identified as significantly over-represented if the Bonferroni-adjusted p-value was < 0.05.

### scBONITA algorithm for development of discrete-state models of pathways

#### Network topologies

ScBONITA infers Boolean regulatory rules/ logic gates for directed networks wherein nodes represent genes and edges represent the regulatory relationships between those genes. These networks contain edge annotations denoting activation/inhibition relationships between nodes, which are exploited by scBONITA to restrict the search space for rule inference to sign-compatible canalyzing functions. Such network topologies of biological pathways are commonly obtained from pathway databases such as KEGG and WikiPathways (142–144). ScBONITA offers an interface to KEGG and WikiPathways databases that allows automated download and processing of user-specified networks. Users can also provide custom networks in graphml format.

#### Boolean rule determination from scRNAseq data

scBONITA assumes that cross-sectional measurements of cells by scRNA-seq data represent steady state of an underlying dynamic biological process. scBONITA’s rule determination (scBONITA-RD) algorithm, which has been extended from our previous BONITA algorithm exploits this property to infer Boolean rules for an input biological network, using a combination of a genetic algorithm (GA) and a node-wise local search (138).

The global search using GA infer a single candidate rule set that adequately describes the input data with respect to the network topology with minimum error (35, 145). The function to be minimized is:

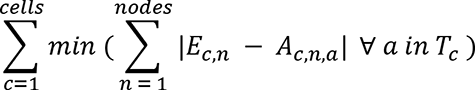

Where *c* from *1* to *cells* iterates over the number of cells in the training dataset, n iterates from 1 to number of *nodes* in the network, *E_c,n_* is the binarized expression of node *n* in cell *c*, *A_c,n,a_* is the value of node *n* in the attractor *a* reachable from cell *c*, and *T_c_* is the attractor reachable from *c*. Note that *T_c_* may have multiple repeating states in a limit cycle or only one steady state, i.e., it may be a singleton attractor. *T_c_* is obtained after simulating the network with the candidate rule set for 100 time-steps, which causes the simulation to reach an attractor state for all tested networks.

The minimum error rule set identified using the above-described genetic algorithm strategy is further refined by a node-level local search that sequentially optimizes the rule for each node keeping the rules for all other nodes in the network constant. An optimal set of rules for a node *n* is obtained by minimizing the function

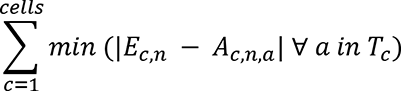

where variables and constants are same as described above.

Several rules may satisfy the termination criteria. The local search returns a set of equivalent rules that all satisfactorily explain the observed state in the experimental data. This set of rules is referred to as the equivalent rule set (ERS) in the text.

#### Pathway analysis (PA) with scBONITA

scBONITA performs pathway analysis in a two-step process. In the first step, importance scores for each node in the biological network under consideration are calculated. In the second step, a pathway modulation metric incorporating both experiment-specific fold changes and the node importance scores calculated in step 1 is calculated.

scBONITA quantifies the influence *I*_*n*_ of a node *n* over the state of the network by quantifying the overall effect of its perturbation on that network. This is achieved by simulating knock-in and knock-out of that node.

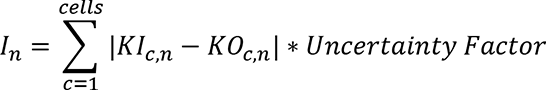

where *KI_c,_*_n_ and *KO_c,n_* are the discrete expression vectors of network node *n* in the attractors reached after a simulation starting from cell *c* where the node under consideration *n* is knocked in and knocked out respectively. The uncertainty factor is defined as follows:

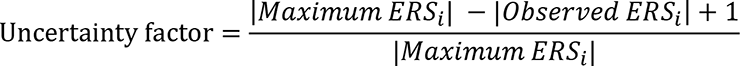

Where *ERS*_*i*_ is the ERS for a node *i*, |*Maximum ERS*_*i*_ | is the maximum possible size of the ERS for a node *i* and |*Observed ERS*_*i*_| is the size of the ERS for a node *i* upon optimization by scBONITA.

The uncertainty factor weighs *I*_*n*_ relative to the maximum state space for that node, to capture the uncertainty in the rule determination for that node. The importance scores of the nodes in a network are scaled to *[0, 1]* by dividing by the maximum calculated importance score for the network under consideration.

A pathway modulation metric (*M*_*p*_) is calculated by weighting the node importance score by the difference between the average gene expression in each group (relative abundance, *RA*) and the standard deviation of expression of that gene (*σ*) across cells. A p-value is calculated by bootstrapping, where a contrast-specific distribution of weighted importance scores is generated using randomly resampled *RA* values. Pathways are described in the text as being overall upregulated in a given contrast if the sum of fold changes of all genes in the pathway is positive. Conversely, pathways are described as being downregulated if the sum of fold changes of all genes in the pathway is negative.

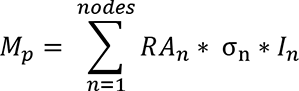

#### Steady-state analysis with scBONITA

scBONITA assumes that the observed cellular states are defined by states of multiple dynamic cellular processes or signaling pathways. While observed cells are samples along a dynamic trajectory of signaling cascades, analyzing attractors upon randomly sampling the rules from ERS allows us to investigate most common signaling states of a network under consideration. We sample ten network specific from the ERS inferred by scBONITA-RD to identify a set of reachable attractors. This is achieved by simulating the network synchronously as performed in other studies (107, 146–148) starting from an observed state (i.e., a cell expression vector) until a steady state (or an attractor cycle) is reached. By starting simulations from observed expression levels of all cells (i.e., all observed states), we can ensure that these simulations cover a large fraction of available state space for a given network. In this way, all reachable attractor states, corresponding to observable signaling states, can be identified. The similarity between cells and attractors is quantified using the Hamming distance. Cells are assigned to the attractor that is closest to their expression data.

#### Implementation and availability

scBONITA is implemented in Python 3 and C. Source code, documentation, and tutorials are available on https://github.com/Thakar-Lab/scBONITA.

### Application of scBONITA on a publicly available data set

A scRNA-seq dataset obtained from four persons living with HIV (PLWH) before and during infection was selected to demonstrate the utility of the scBONITA pipeline on other datasets and to compare signaling dysregulations upon atherosclerosis in PLWH to signaling dysregulations upon HIV infection (38). Log2-transformed TPM data and metadata processed and curated by the study authors was collected from the Single-Cell Portal database (https://singlecell.broadinstitute.org/single_cell/study/SCP256.). The complete scBONITA pipeline was used to compare samples collected before infection to samples collected 1 year after infection. We retained the cluster labels assigned by the authors of the original study. A set of 210 KEGG networks was used with the scBONITA pipeline.

### In silico evaluation of scBONITA

To show that scBONITA-RD is robust to training set size, we selected a cluster of B cells from the HIV/AS dataset. This subset of the dataset was manipulated to either downsample or augment the size of the training dataset (number of cells) presented to scBONITA-RD. The training dataset was downsampled to 1% and 50% of the original number of cells for cluster 0 (“B cells naïve – 1”). To augment the dataset and thereby introduce heterogeneity, the dataset was increased to 200% of its original size by adding in cells from a neighboring cluster of B cells. A set of 210 KEGG networks was used to evaluate the sizes of the ERS obtained by scBONITA-RD using these manipulated training datasets. The size of the ERS is used as a proxy for scBONITA’s ability to successfully cut down the state space of the possible rules for each node using cross-sectional scRNA-seq data.

## Data Availability

The HIV/AS scRNA-seq dataset presented in this manuscript has been deposited in NCBI's Gene Expression Omnibus and will be made accessible through GEO Series accession number GSE198339 on publication of the manuscript. Due to the sensitive nature of HIV data, we have not made the raw data public. All results presented in this manuscript may be recapitulated from the raw count data, processed data, and metadata in GEO. We also analyzed a previously published dataset that is freely accessible at https://singlecell.broadinstitute.org/single_cell/study/SCP256. All source code, tutorials, and documentation for the scBONITA Python package are available at https://github.com/Thakar-Lab/scBONITA.

## List of abbreviations

AS: atherosclerosis
HIV: human immunodeficiency virus
PLWH: people living with HIV
PBMC: peripheral blood mononuclear cell
scBONITA: single-cell Boolean Omics Network Invariant Time Analysis
single-cell RNA sequencing: scRNA-seq
CVD: cardiovascular disease
ORA: overrepresentation analysis
cART: combined antiretroviral therapy
KEGG: Kyoto Encyclopedia of Genes and Genomes
scBONITA-RD: single-cell Boolean Omics Network Invariant Time Analysis rule determination
scBONITA-PA: single-cell Boolean Omics Network Invariant Time Analysis pathway analysis
GA: genetic algorithm
LS: local search
RA: relative abundance
ERS: equivalent rule set
PKN: prior knowledge network

## Availability of data and materials

The HIV/AS scRNA-seq dataset presented in this manuscript has been deposited in NCBI’s Gene Expression Omnibus (149, 150) and is accessible through GEO Series accession number GSE198339 (https://www.ncbi.nlm.nih.gov/geo/query/acc.cgi?acc=GSE198339). Due to the sensitive nature of HIV data, we have not made the raw data public. All results presented in this manuscript may be recapitulated from the raw count data, processed data, and metadata in GEO. We also analyzed a previously published dataset that is freely accessible at https://singlecell.broadinstitute.org/single_cell/study/SCP256. All source code, tutorials, and documentation for the scBONITA Python package are available at https://github.com/Thakar-Lab/scBONITA.

## Article and Author Information

### Authors’ information

#### Authors’ contributions

Conceptualization of the study: JT, conceptualization of BONITA: MGP, RP, JT; Data curation: MGP, AT, JT; Formal Analysis: MGP, JT; Funding acquisition: SM, GS, JT; Investigation: MGP, JT; Methodology: MGP, JT; Project administration: AT, JT; Resources: SM, GS, MVS, JT; Software: MGP, JT; Supervision: JT; Validation: MGP, JT; Visualization: MGP, JT; Writing – original draft: MGP, JT; Writing – review & editing: All authors

## Acknowledgements

We would like to thank Alan Grossfield, Andrew McDavid, Gourab Ghoshal, and all past and present members of the Thakar Lab for helpful discussions on scBONITA implementation and functionality. The Center for Integrated Research Computing at the University of Rochester provided high-performance computing resources and computing expertise. We are especially grateful to the participants in the HIV/AS study, their families, and the clinical team.

## Ethics Declarations

All methods were carried out in accordance with University of Rochester guidelines and regulations, and all experimental and study protocols were approved by the University of Rochester Institutional Review Board (#RSRB00063845). The project does not qualify as human subjects research (45 CFR 46.102) in that the activities do not involve human subjects as defined in the federal regulations because this project utilizes anonymous information.

## Supplementary Material

1. **Supplementary File 1:**

- **Filename:** supplementary_file_1.pdf
- **Description:** Contains Supplementary Note 1 and Supplementary Figures 1 – 7 and corresponding captions.

i. **Supplementary Note 1:** De-identified subject information and description of subject matching in our HIV-associated atherosclerosis study.
ii. **Supplementary Figure 1:** Description of subjects in HIV-associated atherosclerosis (AS) study. There were no significant differences in age, cholesterol levels, high-density lipids (HDL) levels or CD4+ T cell counts between subjects with and without atherosclerosis (two-sided t-test, p > 0.05 for all three comparisons). (A) Distribution of serum lipids (cholesterol and HDL in mg/dl) for subjects with (AS+) and without (AS-) atherosclerosis. (B.) Distribution of CD4+ T cells/µl for subjects with (AS+) and without (AS-) atherosclerosis. (C.) Table showing age, cholesterol, and HDL for each subject.
iii. **Supplementary Figure 2:** Cluster labels were assigned using CIBERSORT in conjunction with a reference set of 22 immune cell types. The 16 clusters are identified on the x-axis as 0 – 15. Each bar represents the proportions of the immune cell types in each cluster.
iv. **Supplementary Figure 3:** Deconvolution of a bulk RNA-seq dataset from persons with and without AS to show the proportion of immune cell types. A bulk RNA-seq dataset obtained from matched PLWH with and without AS was deconvoluted using CIBERSORT to quantify the abundance of the cell subpopulations in the scRNA-seq dataset. Subpopulation-level differences in the percentage of sequenced cells from a subject corresponding to each cell type in Figure 1A (main text) between AS+ and AS- PLWH are identified using a two-sided t-test. The mean of each group is represented by a red asterisk. The population ‘T cells CD8 NK cells resting’ was significantly more abundant in AS- PLWH (t-test, p < 0.1).
v. **Supplementary Figure 4:** Correlation between importance scores for networks trained on subpopulations of PBMCs from the Kazer et al dataset and trained on the corresponding subpopulations from the HIV/AS dataset. (A) T cells from the Kazer et al dataset and subpopulations containing CD4+ T cells from the HIV/AS dataset. (B) B cells from the Kazer et al dataset and subpopulations containing B cells from the HIV/AS dataset. (C) Monocytes from the Kazer et al dataset and from the HIV/AS dataset.
vi. **Supplementary Figure 5:** (A) Distribution of ERS sizes for 130 KEGG networks trained on PBMCs from subjects prior to infection and 1 year after HIV infection (Kazer et al). (B) Distribution of ERS sizes for 130 KEGG networks trained on PBMCs from HIV+ subjects with and without atherosclerosis.
vii. **Supplementary Figure 6:** scBONITA infers biologically meaningful dysregulated (Bonferroni-adjusted p-value < 0.1) pathways for subpopulations of PBMCs derived from HIV-subjects and subjects after 1 year of HIV infection (Kazer et al). Panels A – E show the pathways dysregulated in the HIV-vs 1 year post-infection contrast for clusters labeled as (A) ‘Plasmablast’, (B) ‘Dendritic Cell, (C) ‘Natural Killer Cell, (D) ‘T Cell’ and (E) ‘B Cell’ subpopulations.
viii. **Supplementary Figure 7.** Comparison of genes upregulated in dysregulated pathways (as identified by scBONITA) in the context of HIV-associated atherosclerosis and HIV infection (A) Number of genes upregulated in monocytes derived from AS+ PLWH and in subjects after HIV infection. (B) KEGG gene sets overrepresented in these genes (enrichr, top 10 gene sets shown) (C) Number of genes upregulated in CD8+ T cells derived from AS+ PLWH and in subjects after HIV infection. (D) KEGG gene sets overrepresented in these genes (enrichr, top 10 gene sets shown).
2. **Supplementary Table 1:**

- Filename: *supplementary_table_1.txt*
- **Description of Supplementary Table 1:** Cluster markers for each subpopulation identified in PBMCs derived from AS+ and AS- PLWH are listed in the file “*supplementary_table_1.csv*”. Cluster markers were identified using methods implemented in the Seurat R package, as described in the methods.
3. **Supplementary Table 2:**

- Filename: supplementary_table_2.xlsx
- Description of supplementary table 2: supplementary_table_2 contains 2 worksheets, “DE genes in AS+ vs AS-” and “enrichr_kegg”. “DE genes in AS+ vs AS-” contains a table of genes differentially expressed between cells derived from AS+ and AS- PLWH for each subpopulation in the HIV/AS dataset. The sheet “enrichr_kegg” contains a table of KEGG gene sets enriched (identified using the enrichr R package) in the DE genes from “DE genes in AS+ vs AS-“.
4. **Supplementary Table 3:**

- **Filename:** *supplementary_table_3.txt*
- **Description of Supplementary Table 3:** scBONITA infers biologically meaningful dysregulated pathways for subpopulations of PBMCs derived from AS+ and AS- PLWH in the HIVAS/HIVAS-contrast. The table of dysregulated pathways identified by scBONITA in the HIV+/AS+ - HIV+/AS- contrast in all cell clusters is presented in the CSV file titled “*supplementary_table_3.txt*”.
5. **Supplementary Table 4**

- **Filename:** *supplementary_table_4.txt*
- **Description:** Pearson correlation coefficients between importance scores for networks trained on subpopulations of PBMCs from the Kazer et al dataset and trained on the corresponding subpopulations from the HIV/AS dataset. All p values are < 0.01.
6. **Supplementary Table 5**

- **Filename:** *supplementary_table_5.txt*
- **Description:** scBONITA infers biologically meaningful dysregulated pathways for subpopulations of PBMCs derived from HIV-subjects and subjects after 1 year of HIV infection (Kazer et al). The CSV file *supplementary_table_5.txt* lists the dysregulated pathways and p-values from scBONITA for every subpopulation.
7. **Supplementary Table 6**

- **Filename:** *supplementary_table_6.txt*
- **Description:** Comparison of dysregulated pathways, as identified by scBONITA, between subpopulations of PBMCs derived from HIV-subjects and subjects after 1 year of HIV infection (Kazer et al) and subpopulations of PBMCs derived from HIV+ subjects with and without atherosclerosis. The file *supplementary_table_6.txt* lists the dysregulated pathways for subpopulations from the Kazer et al dataset and the corresponding subpopulations from the HIV/AS dataset, along with the intersections between the pathways dysregulated in the two contrasts. We speculate that intersecting pathways and the pathways dysregulated in the HIV/AS contrast are driven by HIV-associated inflammatory processes. Similarly, pathways that are dysregulated only in the HIV-/HIV+ contrast are assumed to be driven by the immediate antiviral response to HIV infection.

