## Supplementary File 1 for "Executable models of immune signaling pathways in HIV-associated atherosclerosis"

**Supplementary Note 1:**

The association between HIV and atherosclerosis is well-studied by our lab and others (Cornwell et al., 2021; Drozd et al., 2017; Freiberg et al., 2013; Triant, Lee, Hadigan, & Grinspoon, 2007). We recruited a cohort of persons living with HIV with (AS+) and without (AS-) atherosclerosis who were matched as far as possible for the major risk factors for atherosclerosis which would otherwise confound our conclusions. Specifically, we considered the following risk factors. We note that releasing precise values for some of these metrics would constitute a violation of patient privacy and hence we present only generic statements permitted by ethics guidelines.

- Lipid profile (cholesterol, HDL):
  - There were no significant differences between AS+ and AS- subjects based on cholesterol or HDL (t-test, p > 0.05) (Supplementary Figure 1).
- CD4+ T cells/µl:
  - There was no significant difference between AS+ and AS- subjects based on CD4+ T cells/µl (t-test, p > 0.05) (Supplementary Figure 1).
- Smoking:
  - All subjects except one AS- subject are either current or former smokers. One AS+ subject and one AS- subject are current smokers.
  - There was no significant difference between the number of current smokers in the AS+ and AS- groups (chi-squared test, p = 0.5).
- Hypertension status
  - Hypertension:
    - AS+: 1 subject
    - AS-: 2 subjects
  - Hypertension medication:
    - AS+: 2 subjects
    - AS-: 1 subject
- Years of HIV infection:
  - All subjects were infected with HIV for at least 8 years, for an average of ~20 years. There were no significant differences between persons with and without atherosclerosis (two-tailed t-test, p > 0.5).
- HIV treatment:
  - All subjects were on cART for at least 1 year before the study. A previous study has found that inflammation markers in a cohort of 50 HIV+ persons after 1 year of cART were comparable to those of matched healthy individuals (van den Dries, Claassen, Groothuismink, van Gorp, & Boonstra, 2017).

| 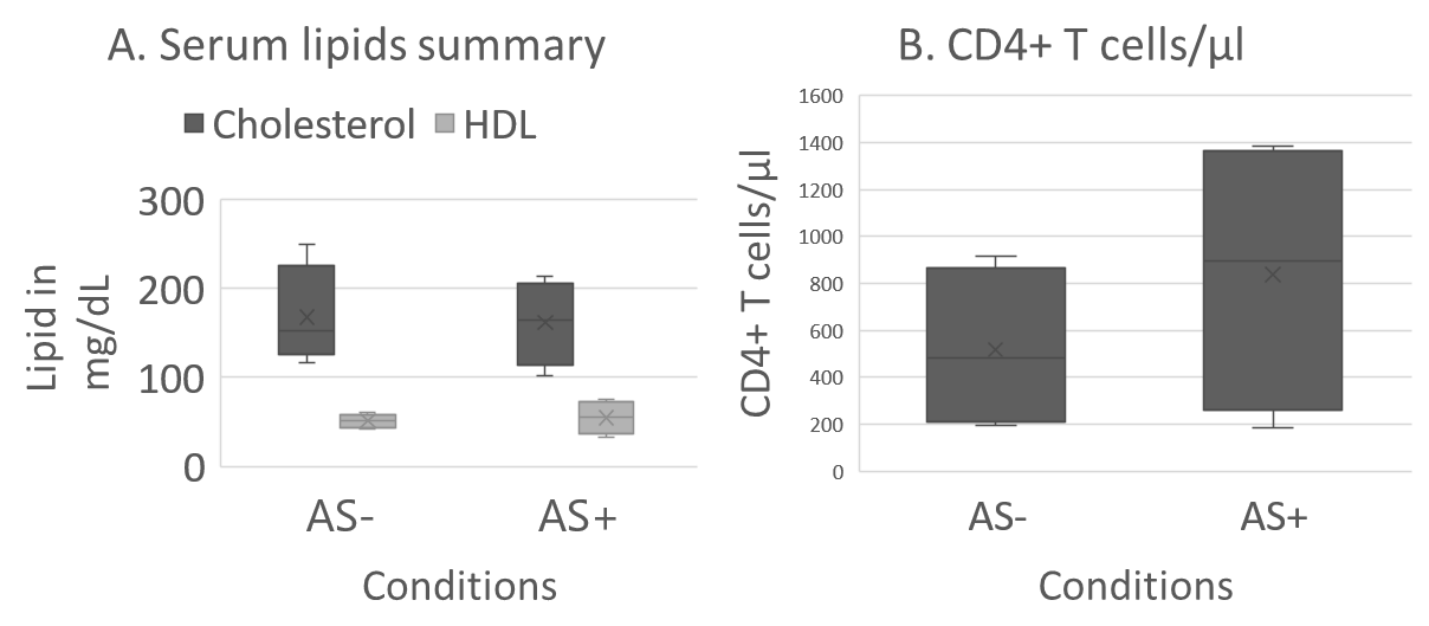  C. | | | | |  |
| --- | --- | --- | --- | --- | --- |
| Subject ID | Condition | Age in years | Cholesterol in mg/dl | HDL in mg/dl | CD4+ T cells/µl |
| Participant_1 | AS- | 41-50 | 151 | 60 | 701 |
| Participant_2 | AS- | 51-60 | 153 | 48 | 195 |
| Participant_3 | AS- | 51-60 | 250 | 42 | 917 |
| Participant_4 | AS- | 51-60 | 117 | 54 | 261 |
| Participant_5 | AS+ | 51-60 | 179 | 65 | 1384 |
| Participant_6 | AS+ | 51-60 | 102 | 33 | 182 |
| Participant_7 | AS+ | 51-60 | 151 | 46 | 1297 |
| Participant_8 | AS+ | 61-70 | 214 | 75 | 492 |

**Supplementary Figure 1:** Description of subjects in HIV-associated atherosclerosis (AS) study. There were no significant differences in age, cholesterol levels, high-density lipids (HDL) levels or CD4+ T cell counts between subjects with and without atherosclerosis (two-sided, p > 0.05 for all three comparisons). (A) Distribution of serum lipids (cholesterol and HDL in mg/dl) for subjects with (AS+) and without (AS-) atherosclerosis. (B.) Distribution of CD4+ T cells/µl for subjects with (AS+) and without (AS-) atherosclerosis. (C.) Table showing age, cholesterol, and HDL for each subject.

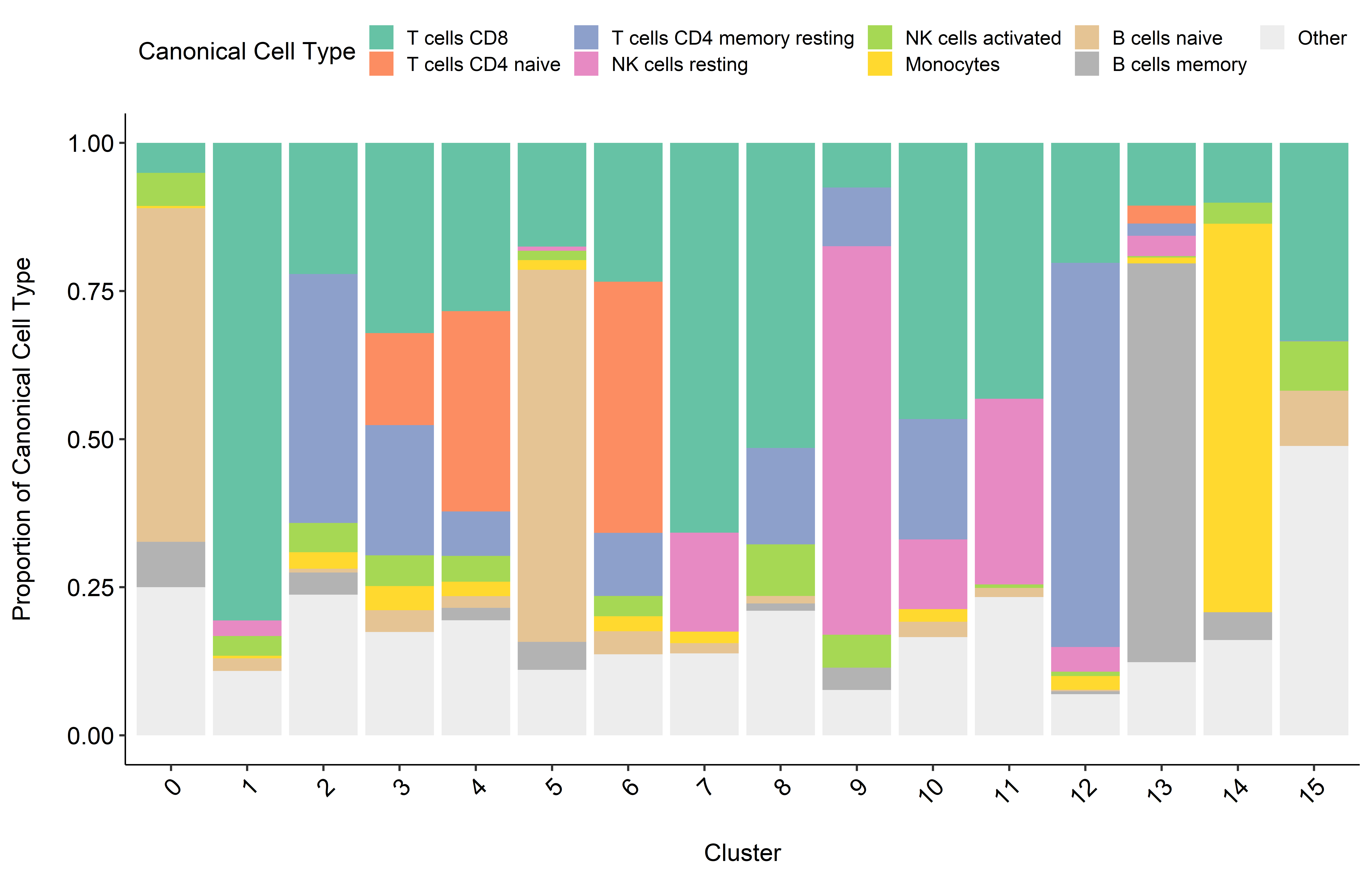

**Supplementary Figure 2:** Cluster labels were assigned using CIBERSORT in conjunction with a reference set of 22 immune cell types. The 16 clusters are identified on the x-axis as 0 – 15. Each bar represents the proportions of the immune cell types in each cluster.

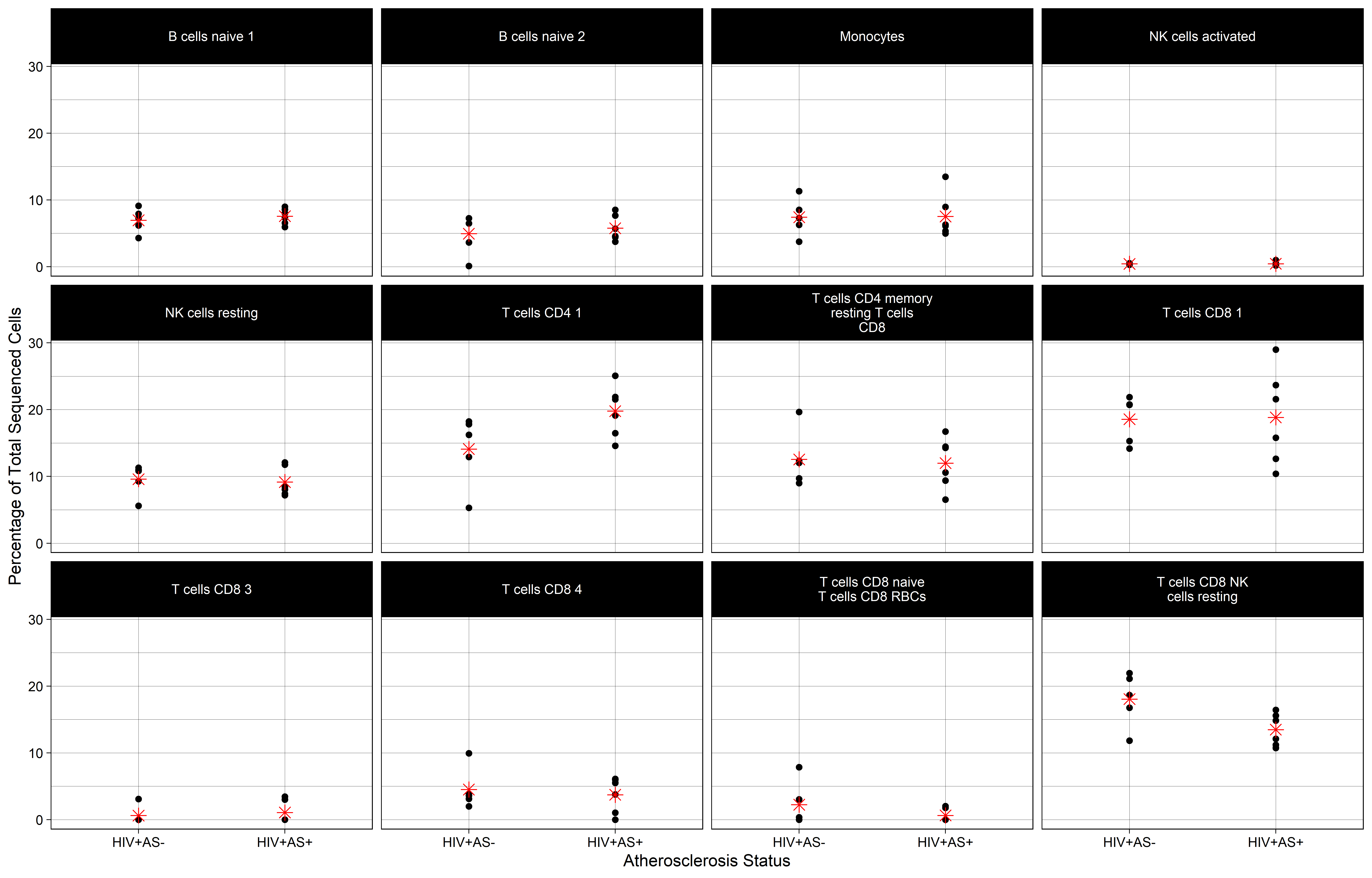

**Supplementary Figure 3: Deconvolution of a bulk RNA-seq dataset from persons with and without AS to show the proportion of immune cell types.** A bulk RNA-seq dataset obtained from matched PLWH with and without AS was deconvoluted using CIBERSORT to quantify the abundance of the cell subpopulations in the scRNA-seq dataset. Subpopulation-level differences in the percentage of sequenced cells from a subject corresponding to each cell type in Figure 1A (main text) between AS+ and AS- PLWH are identified using a t-test. The mean of each group is represented by a red asterisk. The population ‘T cells CD8 NK cells resting’ was significantly more abundant in AS- PLWH (t-test, p < 0.1).

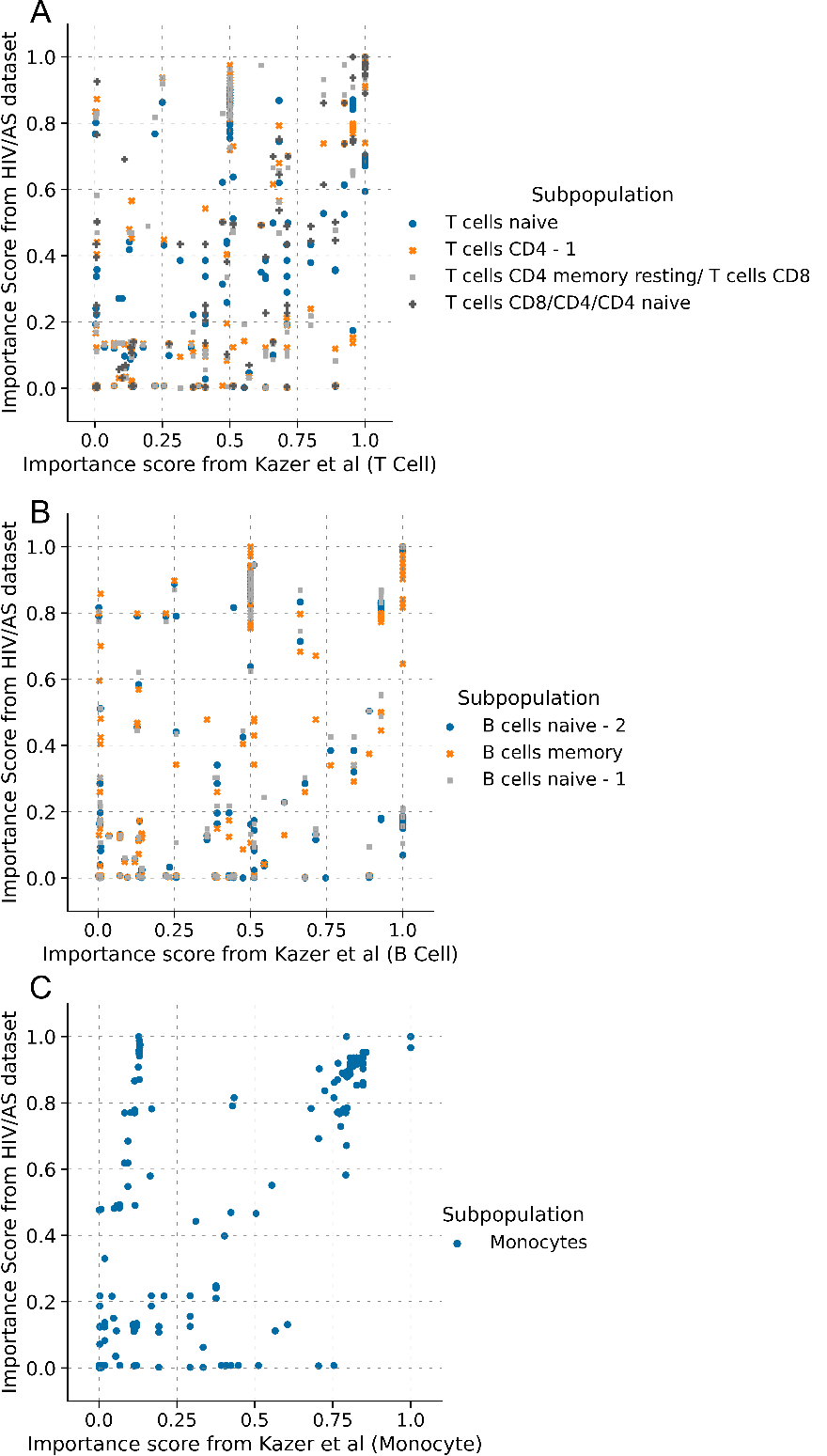
**Supplementary Figure 4:** Correlation between importance scores for networks trained on subpopulations of PBMCs from the Kazer et al dataset and trained on the corresponding subpopulations from the HIV/AS dataset. (A) T cells from the Kazer et al dataset and subpopulations containing CD4+ T cells from the HIV/AS dataset. (B) B cells from the Kazer et al dataset and subpopulations containing B cells from the HIV/AS dataset. (C) Monocytes from the Kazer et al dataset and from the HIV/AS dataset.

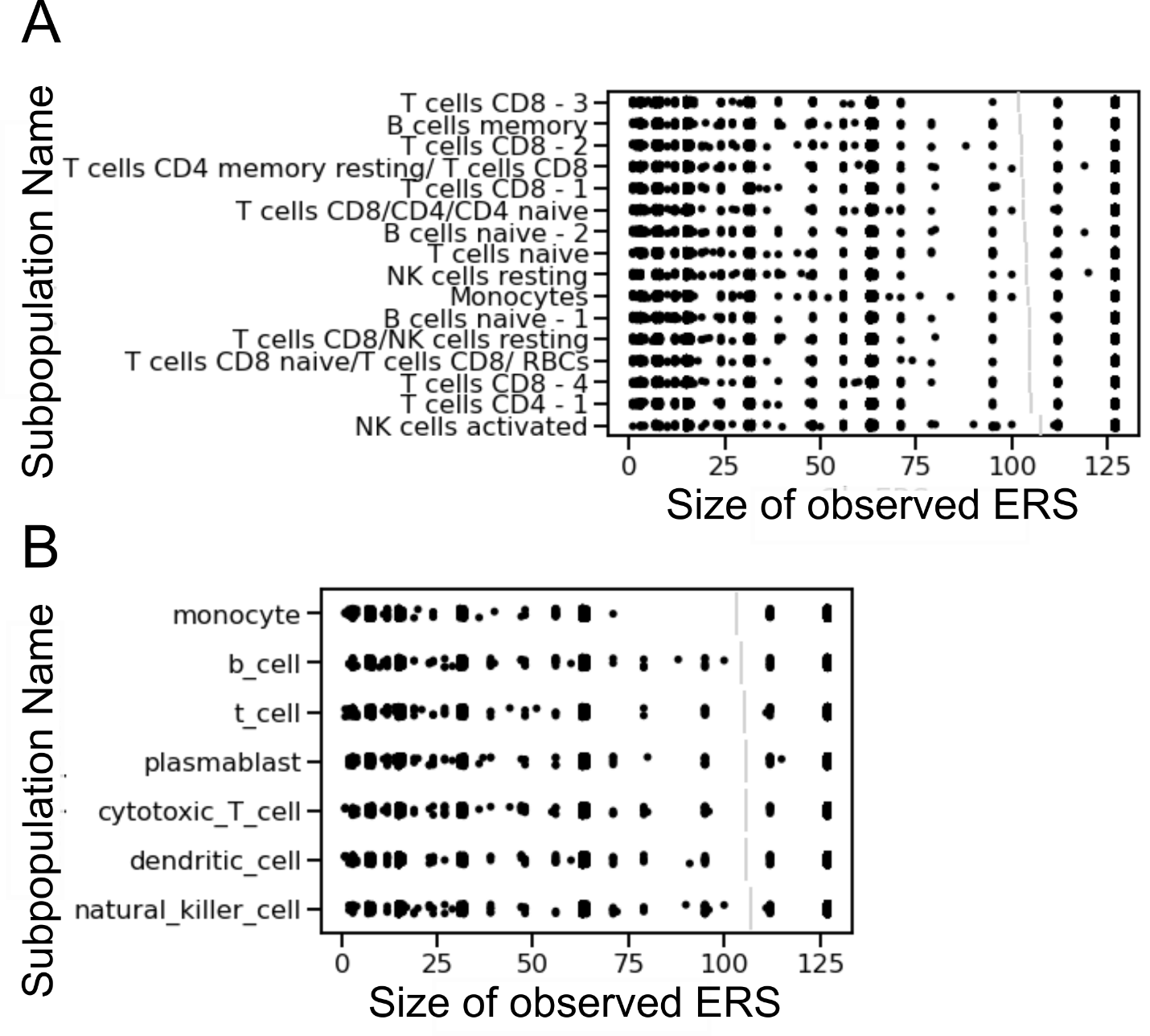

**Supplementary Figure 5:** (A) Distribution of ERS sizes for 130 KEGG networks trained on PBMCs from subjects prior to infection and 1 year after HIV infection (Kazer et al). (B) Distribution of ERS sizes for 130 KEGG networks trained on PBMCs from HIV+ subjects with and without atherosclerosis.

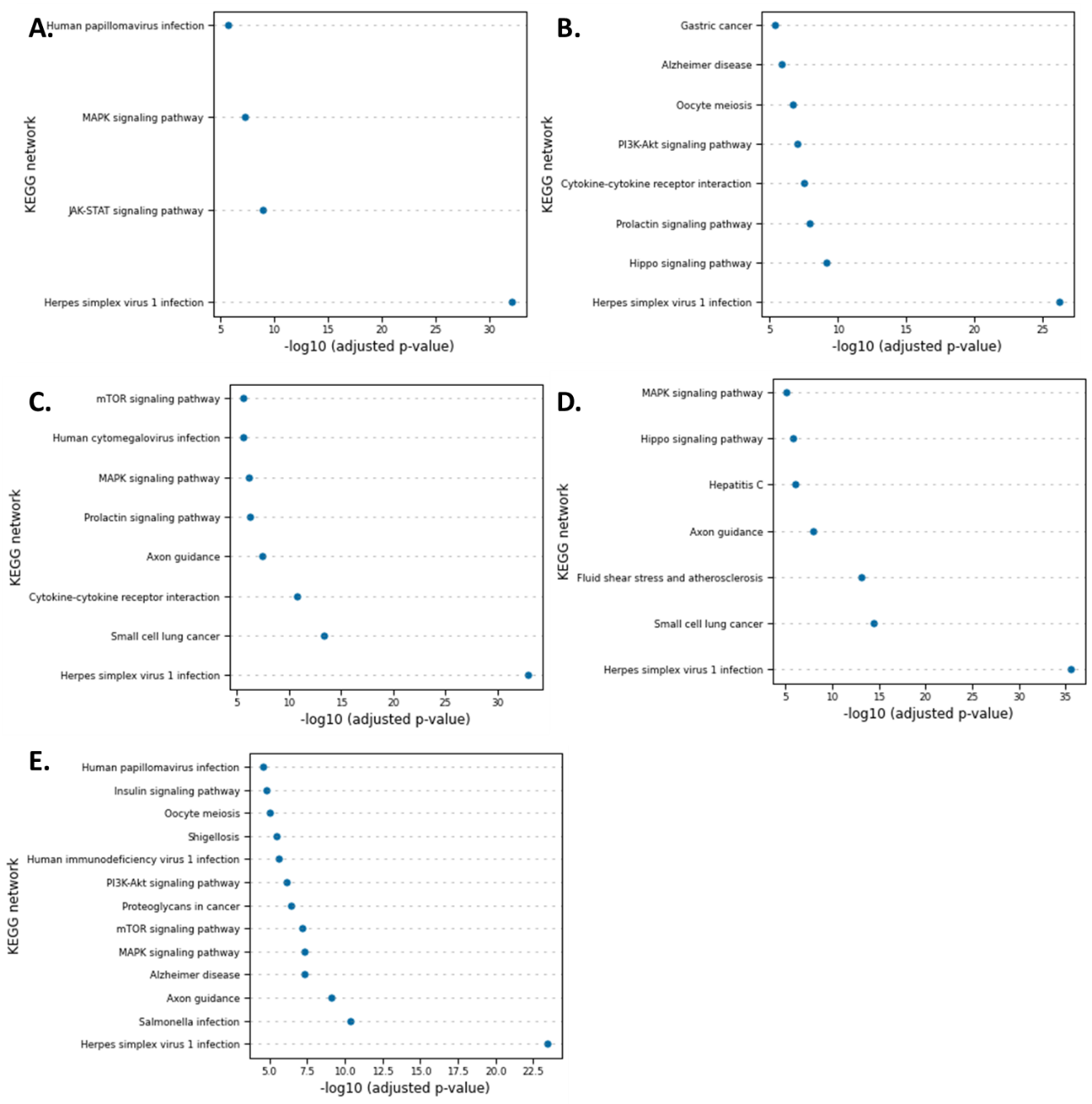

**Supplementary Figure 6:** scBONITA infers biologically meaningful dysregulated (Bonferroni-adjusted p-value < 0.1) pathways for subpopulations of PBMCs derived from HIV- subjects and subjects after 1 year of HIV infection (Kazer et al). Panels A – E show the pathways dysregulated in the HIV- vs 1 year post-infection contrast for clusters labeled as (A) 'Plasmablast’, (B) 'Dendritic Cell, (C) 'Natural Killer Cell, (D) 'T Cell’ and (E) ‘B Cell’ subpopulations.

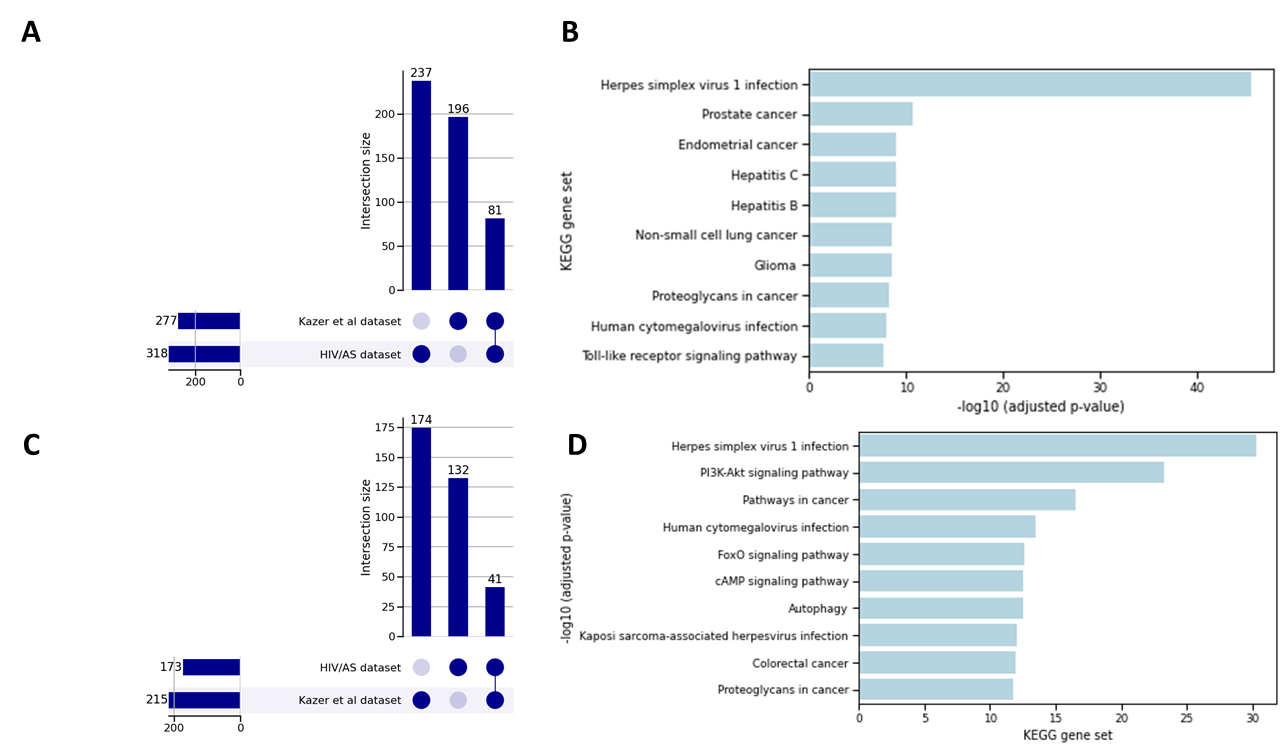

**Supplementary Figure 7.** Comparison of genes upregulated in dysregulated pathways (as identified by scBONITA) in the context of HIV-associated atherosclerosis and HIV infection (A) Number of genes upregulated in monocytes derived from AS+ PLWH and in subjects after HIV infection. (B) KEGG gene sets overrepresented in these genes (enrichr, top 10 gene sets shown) (C ) Number of genes upregulated in CD8+ T cells derived from AS+ PLWH and in subjects after HIV infection. (D) KEGG gene sets overrepresented in these genes (enrichr, top 10 gene sets shown).
